# Conserved roles of cardiac endothelial ETS factors in ventricular development and disease

**DOI:** 10.64898/2026.09.08.26361286

**Authors:** Lu Wang, Analyne Schroeder, Katya Marchetti, Ingolf Reim, Matthias Blanc, K’leigh Guillotte, Amelie Mück, Elaina Bao, Zhi Ling, Shu Jia, Denise Malicki, Rolf Bodmer, Shuyi Nie, Karen Ocorr, Paul Grossfeld

## Abstract

We previously identified ETS1 as a candidate gene in the rare chromosomal deletion disorder, Jacobsen syndrome (JS, OMIM#147791). Here, we present data from four model systems that define roles for ETS factors in normal heart development and suggest how defects in these genes contribute to congenital heart disease. In frog, knockdown of *Ets1* in cardiac mesoderm led to a hypoplastic ventricle with impaired cardiac function and loss of trabecular myocardium. Grafting of wildtype cardiac mesodermal tissue restored the integrity of the ventricular endocardial layer and normal development. In zebrafish, KD of *ETS1* and two related *ETS* factor genes caused severely underdeveloped, poorly functioning ventricles with decreased numbers of endocardial cells that failed to contact the myocardium. Genetic ablation of the endocardium in mice also caused a hypoplastic ventricle with loss of the trabecular myocardium. In *Drosophila*, loss of the *ETS1* ortholog *pointed* (*pnt*) affected specification of cardiac precursors and decreased Notch signaling, a pathway previously implicated in some forms of hypoplastic left heart syndrome (HLHS). We also demonstrated genetic interactions between *pnt* and *tinman*, the *NKX2.5* ortholog, associated with HLHS. Finally, our examination of the heart from a newborn with JS/HLHS showed myofibrillar disarray and myocardial maturation defects. Furthermore, there was a reduction in the coronary vascular endothelium in the LV myocardium from the JS/HLHS patient compared to an age-matched normal heart. Taken together, our studies demonstrate a critical and conserved role of ETS factors for cardiac endothelial function and in ventricular morphogenesis.

## Introduction

Jacobsen Syndrome (JS, OMIM #147791) is a rare chromosomal disorder caused by deletions within the long arm of chromosome 11 ^1,2^. About half of affected infants have structural heart defects, most commonly septal and left-sided obstructive defects, including hypoplastic left heart syndrome (HLHS, Figure 1 B-E), which is 200 times more frequent in JS than in the general population. Our previous studies led to the identification of a “cardiac critical region” in 11q containing six annotated genes including ETS1 and FLI1, both members of the ETS family of transcription factors ^3^. We have previously identified a patient with a complex congenital heart defect, including a hypoplastic left ventricle, carrying a *de novo* loss of function mutation in ETS1 ^4^.

**Figure 1.**
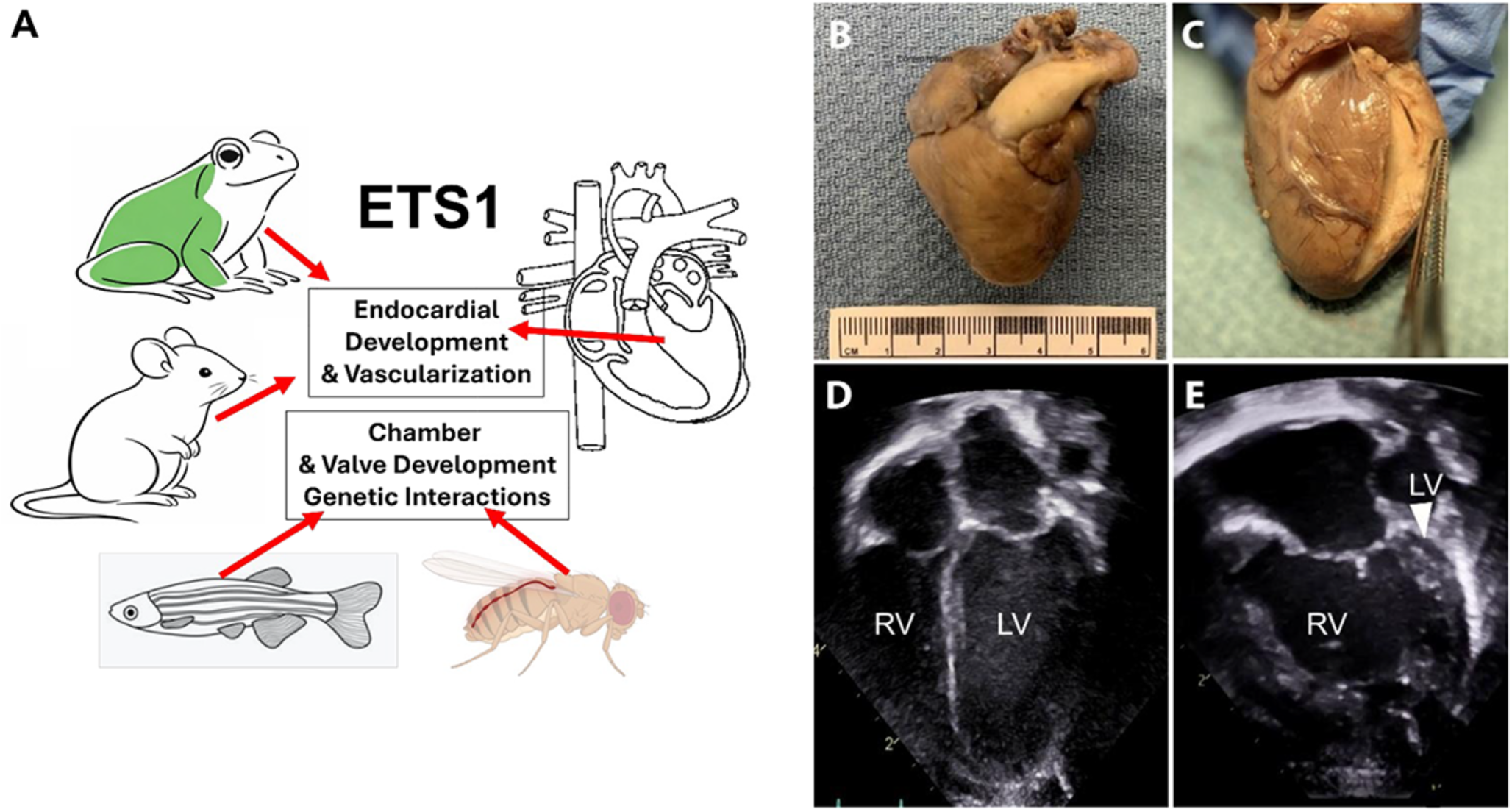
**(A)** Graphical Abstract. **(B)** Intact heart from the HLHS patient with Jacobsen syndrome (14 Mb terminal deletion in 11q) showing a severely dilated pulmonary artery. **(C)** Dissected heart showing the slit-like left ventricular cavity (arrow). **(D)** Subcostal echocardiographic view of a normal human heart from a female infant. **(E)** Subcostal echocardiographic view of a newborn JS/HLHS patient showing a slit-like left ventricle (arrow) with an atretic mitral valve.

In mouse, ETS1 is expressed in the embryonic neural crest and cardiac endothelial lineages. Conditional deletion of ETS1 in the neural crest leads to double outlet right ventricle and a dysplastic aortic valve due to impaired cell migration ^5^. *In vitro* and *in vivo* studies demonstrated impaired neural crest cell migration and increased expression of the adhesion molecule N-Cadherin. Cardiac endothelial deletion causes ventricular non-compaction: an overgrown trabecular layer, and a thinned compact zone. Loss of ETS1 in the cardiac endothelium impairs cell migration and vascular formation, with a reduction of secreted factors that promote myocyte proliferation ^6,7^.

Here we use fish, frog and fruit fly, to delineate the roles of Ets1 signaling and cardiac endothelium in early cardiac development. We observed in frog, zebrafish and adult fruit fly that loss of ETS function disrupts ventricular development, leading to a hypoplastic ventricle. Furthermore, we provide *in vivo* evidence demonstrating that ETS factors play a critical role in the endocardium, contributing to the hypoplastic ventricular phenotype. In *Drosophila*, we identified a strong genetic interaction between *pnt*/*Ets1* and *tinman*/*Nkx2.5*. Finally, analysis of heart tissue from a newborn with JS and HLHS revealed cardiomyocyte maturation defects and reduced coronary microvasculature. Together, these findings support a model in which defects in cardiac endothelial lineages impair normal ventricular growth in a subset of HLHS patients.

## Methods

Materials and Methods are detailed in Supplemental Material.

## Results

### *ETS genes affect* cardiac development in frog

Loss of Ets1 in frog embryos leads to severe heart defects ^8^, predominantly a small ventricle with reduced chamber size, resembling the MA/AA subtype of the human HLHS left ventricle. Using an ultrafast volumetric live imaging technique, Fourier light-field microscopy (FLFM), we directly analyzed heart performance in live tadpoles ^9,10^. To distinguish individual cardiac cells, nuclear GFP (H2b-EGFP) was injected into the lateral medial marginal zone of cleavage stage embryos where prospective cardiogenic mesoderm resides, with or without Ets1 morpholinos. Stage 46 embryonic tadpole hearts were imaged for 10 seconds at 10 ms intervals and significant differences in ventricular function were observed between wildtype and morphant hearts. Control hearts beat in a rhythmic manner and contract efficiently, many morphant hearts only twitched (**Supplemental Movie 1,2**). Heart rate, diastolic and systolic ventricular size (outer diameters) and ventricular chamber size (lumen volume), ejection fraction, and cardiac output were calculated for 9 wildtype and 7 mutant embryos. The end diastolic ventricular size and ventricular chamber size decreased significantly in morphant hearts compared to controls (**Figure 2A-B**). There was also a moderate decrease in the average heart rate but this did not reach the level of significance (**Figure 2C**). While the decreased systolic ventricular size and ventricular chamber size were not statistically significant, ventricular contractility (measured as ejection fraction) was dramatically decreased by Ets1 knockdown (KD, **Figure 2D**). Cardiac output was also significantly decreased (**Figure 2E**).

**Figure 2.**
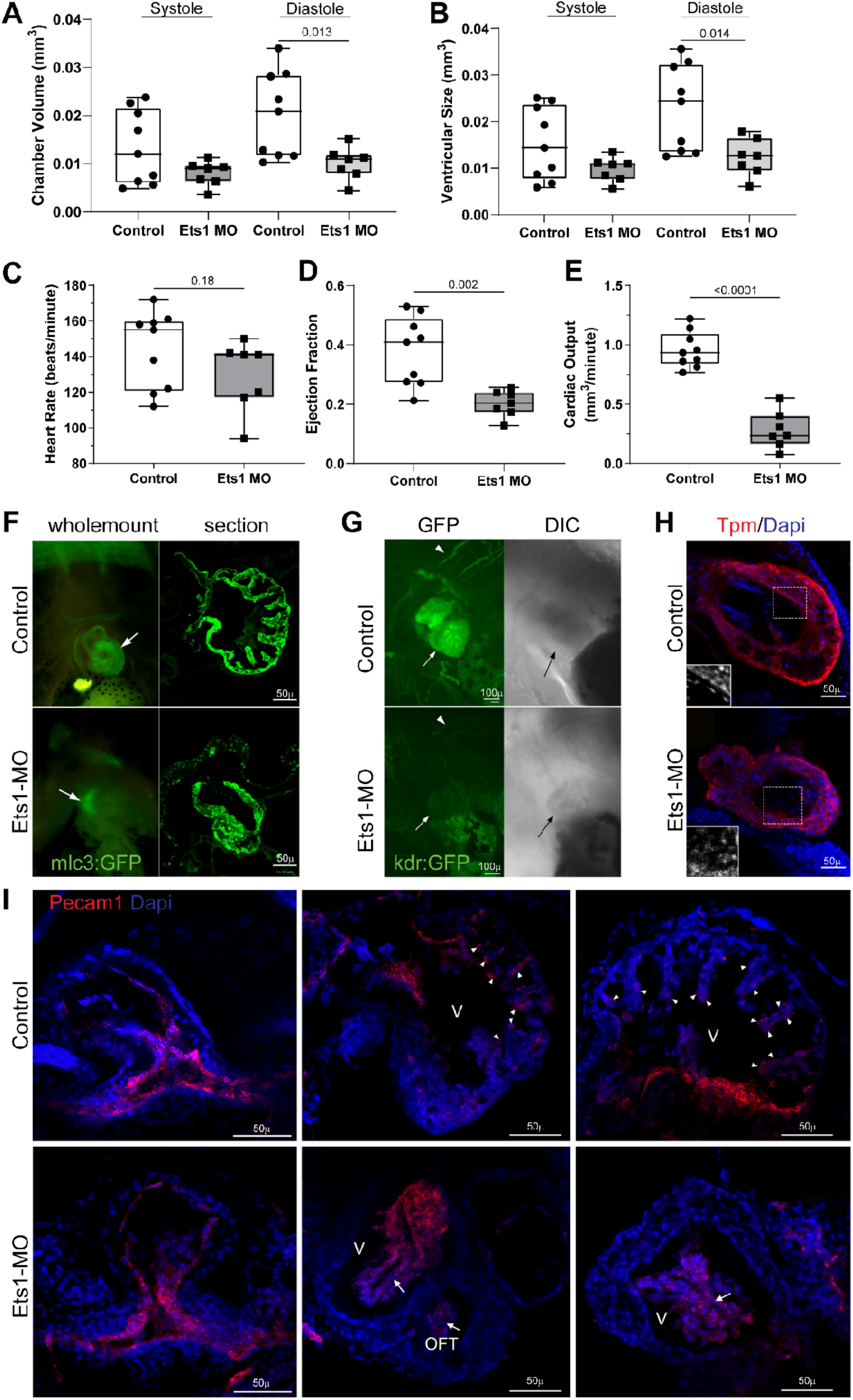
**(A)** Ventricular chamber volume, **(B)** ventricle size, **(C)** heart rate, **(D)** ejection fraction, and **(E)** cardiac output measured by Fourier light field microscopy in wild-type and Ets1 knockdown hearts. Plots show minimum, first quartile, median, third quartile, and maximum. Significance by unpaired Student’s *t*-test. **(F)** Cardiomyocytes in *mlc3:GFP* transgenic tadpoles. **(G)** Endocardial/endothelial cells in *kdr:GFP* tadpoles (left) with DIC image (right). Arrowheads indicate cranial vasculature; arrows indicate cardiac tissue. **(H)** Immunofluorescence for cardiomyocytes (Tropomyosin) and nuclei (DAPI). Endocardial nuclei are adjacent to cardiomyocytes in control ventricles but not in Ets1 knockdown hearts. **(I)** HCR showing *Pecam1* expression in endocardial/endothelial tissue of aortic arch arteries and distal outflow tract (arrowheads). In Ets1 knockdown hearts, *pecam1* signal appears within ventricular chambers (arrows).

To assess tissue organization, we examined myocardial and endocardial lineages in transgenic frogs expressing myl3:GFP (to visualize myocardial cells) and kdr:GFP (to visualize endothelial cells) (National Xenopus Resources ^11^;^12^). Tadpoles were raised to stage 46 (when heart chambers are fully developed), then fixed and optically cleared for wholemount imaging. Ventral view of the wildtype myl3:GFP embryo shows well-organized myocardial cells in the heart, with a thin layer of GFP positive cells lining the atria and GFP-positive cells populating the ventricle (Figure 2F, top). In contrast, there is a much smaller and irregularly shaped myocardial population in Ets1 morphant heart (**Figure 2F**, bottom). Transverse sections through the heart further showed that in wild type ventricles, cardiomyocytes formed well-organized trabecular structures. In contrast, in Ets1 KD ventricles, cardiomyocytes were disorganized and failed to trabeculate. This result suggests that Ets1 is required for myocardial morphogenesis, but not for cardiomyocyte specification. When examining the endocardial lineage, we observed diminished GFP signal in Ets1 morphant hearts. Despite the obvious presence of a (smaller) heart in the DIC image of morphant tadpoles, kdr positive endocardial/endothelial cells could barely be detected (**Figure 2G**, arrows). This suggests that Kdr expression in frog heart is significantly decreased by the loss of Ets1, consistent with the report that kdr is regulated downstream of Ets1 in the vascular system ^13^.

To determine whether endocardial cells were present in these mutant hearts, we combined nuclear DAPI staining with immunofluorescence staining against the cardiomyocyte marker gene tropomyosin (Tpm/CH1). In controls, we observed DAPI labelled nuclei adjacent to CH1-positive ventricular cardiomyocytes before trabeculation occurs (**Figure 2H**). In Ets1 morphant hearts, nuclei are present inside the myocardial tissue, but the arrangement of these nuclei was largely random, suggesting defects in the organization of endocardial cells. We also examined the expression of another endocardial/endothelial gene, pecam1, using in situ hybridization chain reaction (HCR) analysis. Pecam1 transcripts were observed in cells in the inner lining of the outflow tract and aortic arch arteries (**Figure 2 I**, left panels) and the ventricles (**Figure 2 I**, right panels). The development of the aortic arch arteries was relatively normal in Ets1 KD hearts and the expression of pecam1 transcripts in Ets1 KD aortic arch arteries was comparable to its expression in control hearts, indicating that Ets1 loss in the mesoderm does not affect the development of the aortic arch arteries. This also suggests that the cardiac defects we observed in the ventricle do not result from a primary outflow tract defect. In contrast, the expression of pecam1 appears very different in the ventricles of morphant hearts. Instead of being expressed next to trabecular cardiomyocytes in control ventricles (white arrowheads), many pecam1 transcripts were observed in the center of the ventricular chamber (white arrows). DAPI staining showed that these pecam1 expressing nuclei were aggregated without any clear organization. Together, these results demonstrate that Ets1 plays important roles in endocardial lineage development and loss of Ets1 results in both defective expression of endocardial genes and disrupted organization of endocardial tissue.

Our results in frog demonstrated that loss of Ets1 leads to early defects in ventricular morphogenesis. We performed a transplantation experiment to determine whether such an early heart defect can be restored by replacing Ets1 KD tissue with wild type tissue. We grafted wildtype heart field tissue into Ets1 morphant embryos at tailbud stages, before linear heart tube formation. The transplanted embryos were raised to tadpole stages, and their heart development was assessed by immunohistochemistry against the myocardial marker gene MYH1E (MF20, **Supplemental Figure 1A**) or Tropomyosin (CH1, **Supplemental Figure 1B**). Optical sections through the heart show that, while Ets1 morphant hearts have a compact ventricle with disorganized myocardial tissue and a diminutive lumen, the grafted heart has significantly improved anatomy with ventricular sizes comparable to controls and an obvious chamber lumen. DAPI staining showed a row of nuclei lining the myocardial tissue (**Supplemental Figure 1B**), similar to that in control hearts, suggesting that myocardial and endocardial organization as well as lumen formation were restored.

### Heart function is compromised with knockout of ETS genes in zebrafish

We knocked out Ets1 using CRISPR-Cas9 F0 mutagenesis ^14^ and assessed heart function in zebrafish embryos at 72 hours post fertilization (hpf) larvae by high-speed video imaging ^15^. Loss of *Ets1* alone had little effect on heart size but caused reduced contractility, measured as fractional area change (FAC, **Figure 3A**), with slightly increased end-systolic surface area (**Figure 3B**,) but no change in end-diastolic surface area (**Figure 3C**), consistent with systolic dysfunction. In a small number of embryos there was evidence of developmental defects in the atrioventricular canals (AVC) and/or outflow tracts (OFT, 3/20 embryos, **Supplemental Movie 3, Supplemental Table 2**).

**Figure 3.**
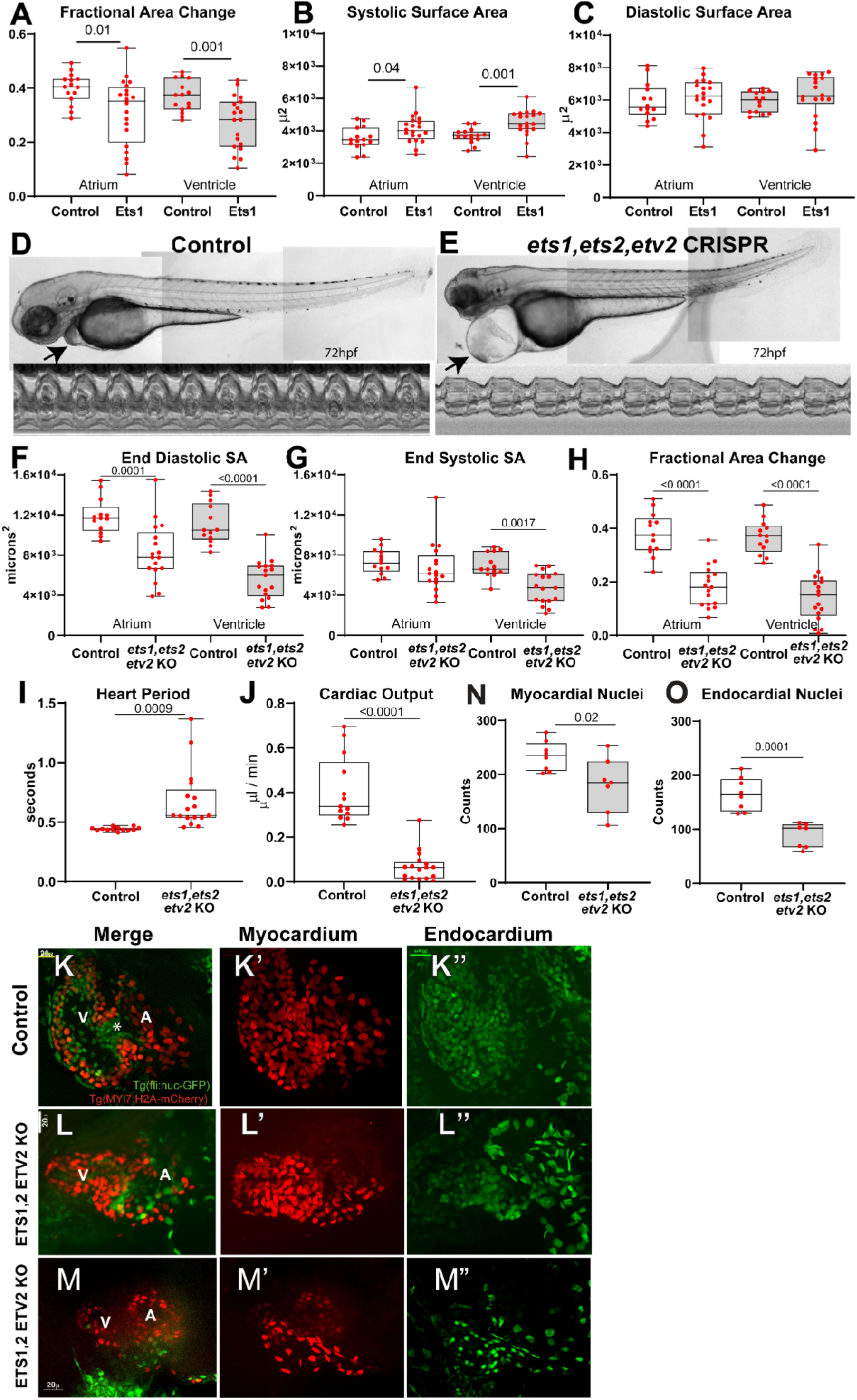
Cardiac function parameters from high-speed movies. **(A)** Fractional Area Change, **(B)** End Systolic surface area, and **(C)** End Diastolic surface area for atria and ventricles from control injected and *ets1* Crisper KO zebrafish embryos (72hpf). **(D)** Control-injected zebrafish with M-mode (5 s), arrows mark the pericardial sac. **(E)** Triple *ets1, ets2, etv2* CRISPR KO embryos with normal body morphology but marked pericardial edema. **(F)** End Diastolic surface area, **(G)** End Systolic surface area, **(H)** Fractional Area Change, **(I)** Heart period (R-R interval) and **(J)** Cardiac Output from Control and triple KO embryos. **(K)** Immunohistochemistry of a control heart expressing fli:nuclear-GFP (endocardium, green) and Myl7:H2A-mCherry (myocardium, red). Optical section shows the ventricular chamber (V), AV canal (*), and atrium (A). **(K’)** Z-stack of Myl7:H2A-mCherry-positive cells. **(K”)** Z-stack of fli:nuclear-GFP-positive nuclei. **(L)** Optical section of a moderately affected crispant heart showing reduced looping. **(L’)** Z-stack of Myl7:H2A-mCherry-positive nuclei. **(L”)** Z-stack of fli:nuclear-GFP-positive nuclei. **(M)** Optical section of a severely affected crispant heart. **(M’)** Z-stack of Myl7:H2A-mCherry-positive nuclei. **(M”)** Z-stack of fli:nuclear-GFP-positive nuclei. **(N)** Quantification of myocardial nuclei and **(O)** endocardial nuclei. **(A-C, F-H)** Significance by one-way ANOVA, Sidak’s multiple-comparisons post hoc test; **(L-O)** significance by unpaired Student’s *t*-test.

In zebrafish, *ets1*, *ets2*, and *etv2* are closely related and may function redundantly ^16–18^. Therefore, we used CRISPR to create triple Ets1,2, Etv2 KO larva. At 72 hpf, F0 mutants had relatively normal body morphology and tail musculature compared with controls (**Figure 3D&EB**). However, triple mutant hearts were more linear, consistent with looping defects, and showed marked pericardial edema, indicating impaired function. Functional analysis showed a significant reduction in End Diastolic and End Systolic surface area as well as reduced contractility (**Figure 3F-H**). All control hearts exhibited identifiable AVC and patent OFT, however in triple ETS factor crispants the AVC appeared only as a constricted mid-heart region and was nonfunctional in about half of the fish, while the OFT was non-patent in most cases (**Supplemental Movie 4, Supplemental Table 2**).

We next examined effects on cardiac cell fate and morphology in 72 hpf double transgenic fish expressing fli:nuclear-GFP (endocardium) and *myl7*:H2A-mCherry (myocardium) ^19^. In controls, immunostaining showed two well-defined looping chambers with a clear AVC, and endocardial cells were closely opposed to myocardial cells (**Figure 3K**). In *ets1/ets2/etv2* KO hearts, chambers were less distinct, looping was reduced, the AVC was poorly defined and myocardial and endocardial layers failed to contact each other (**Figure 3L,M**). Finally, both myocardial and endocardial cell numbers were significantly reduced (**Figure 3N,O**).

### Genetic ablation of the endocardium during early heart development causes ventricular hypoplasia with decreased trabeculation in the mouse heart

To confirm a role for endocardium in ventricular development we created an endocardial ablation model. Nfatc1Cre mice, which express Cre recombinase specifically in endocardial cells, were crossed with Rosa;DTA (Diphtheria toxin fragment A) mice to specifically ablate endocardial cells during murine heart development. (**Figure 4A**). Since Nfatc1 expression initiates around E9, we examined ventricular development starting at E9.5. Initial whole-embryo analysis revealed no obvious morphological differences between Nfatc1Cre;DTA mutant embryos **(Figure 4B2**) and littermate controls (**Figure 4B1**) at E9.5. However, by E10.5, growth retardation and impaired cardiac development became evident in mutant embryos (**Figure 4B4**). Notably, all mutant embryos died before E11.5 (**Figure 4B6**). ERG immunofluorescence staining was performed to visualize the endocardium. Embryos were stained for ERG (marker for endothelial cells), *α*-actinin (marker for cardiomyocytes), and DAPI (nuclei). At E9.5, the morphology of the mutant heart was not significantly different from that of littermate controls (**Figure 4C,D**). However, by E10.5, the mutant heart failed to develop distinct left and right ventricular structures (**Figure 4E,F**). In addition, trabecular development was impaired in the mutant hearts, with the typical finger-like trabecular structures absent (**Figure 4C’, 4D’, 4E’** and **4F’**). Together, these results are consistent with an essential endocardial role in normal ventricular development, as implicated by studies of ETS factor function described above.

**Figure 4.**
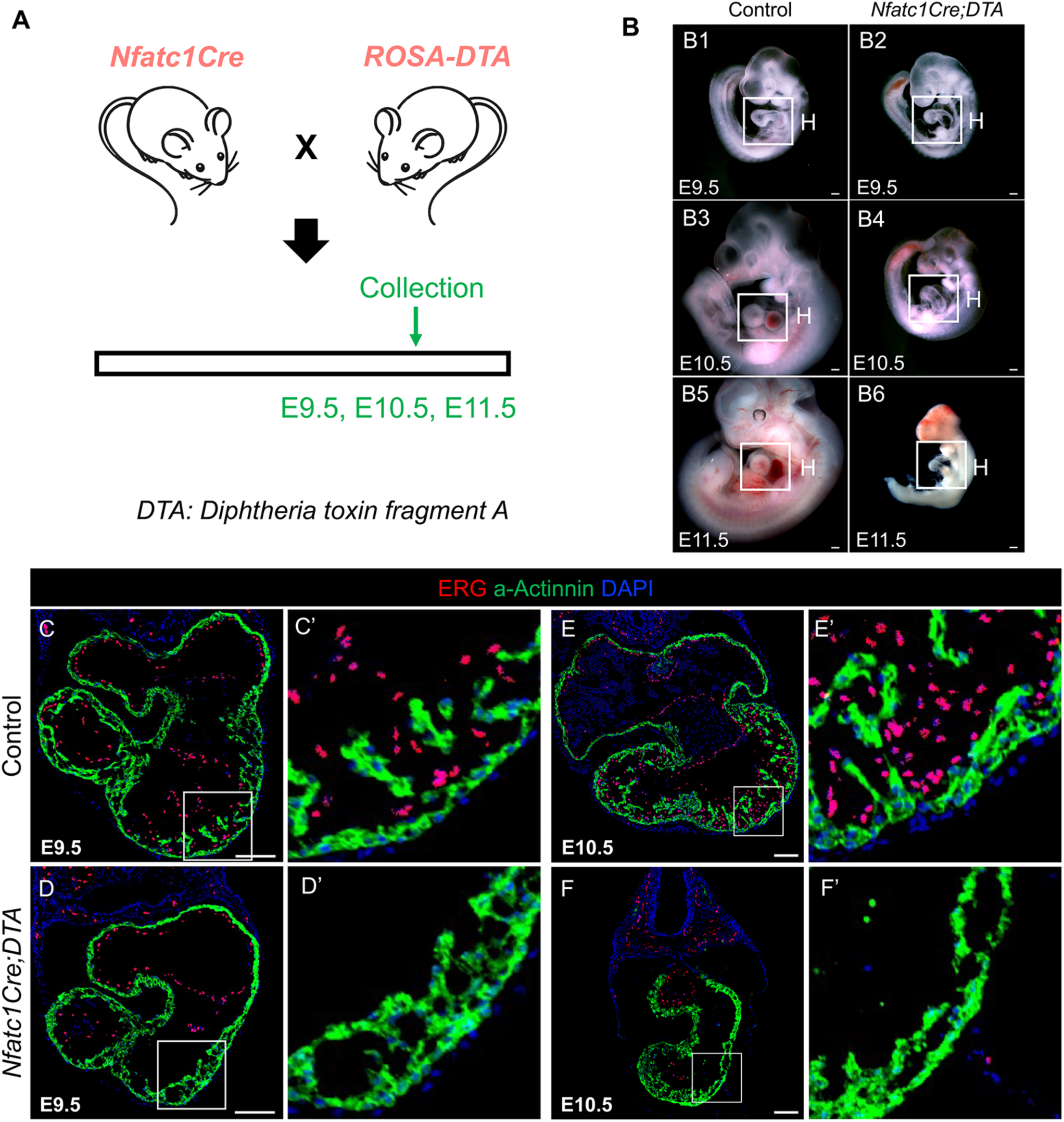
**(A)** Schematic illustrating the endocardial ablation strategy using the Nfatc1Cre driver. **(B)** Representative images of whole embryos from control and Nfatc1Cre;DTA mice at E9.5 **(B1,B2)**, E10.5 **(B3,B4)**, and E11.5 **(B5,B6)**. **(C-F)** Confocal images of ERG (endocardium) and α-actinin (myocardium) immunofluorescence at E9.5 **(C,C’,D,D’)** and E10.5 **(E,E’,F,F’)**. Scale bars, 100 μm.

### Mesoderm-specific knockdown of Ets homolog pnt in Drosophila changes cardiac cell Identity

The single gene *pointed (pnt)* is the *Drosophila* homolog of Ets1. Germline *pnt/Ets1* mutations alter cardioblast and pericardial cell numbers in the embryonic heart, nearly doubling cardioblasts while significantly reducing pericardial cells ^20^. Most of this increase reflects expansion of *sevenup* (*svp*)-positive cardioblasts (∼3-fold), whereas *tinman* (*tin*)-positive cardioblasts increase more modestly (8–20%). Because *pnt* also functions broadly in early development and patterning ^21,22^, we knocked it down specifically in the mesoderm using the GAL4 system ^23^ and quantified changes in cardiac and pericardial cell types (**Figure 5A**). Embryos from *Twist*-GAL4 × UAS-*pnt*-RNAi crosses were stained for *sevenup*, *tinman*, *oddskipped*, and *evenskipped*. Mesodermal *pnt* knockdown caused a milder embryonic phenotype than germline mutation but still expanded *svp*-positive cells in a subset of heart and aortic segments, significantly increasing total *svp*^+^ cell number (**Figure 5B,C**).

**Figure 5.**
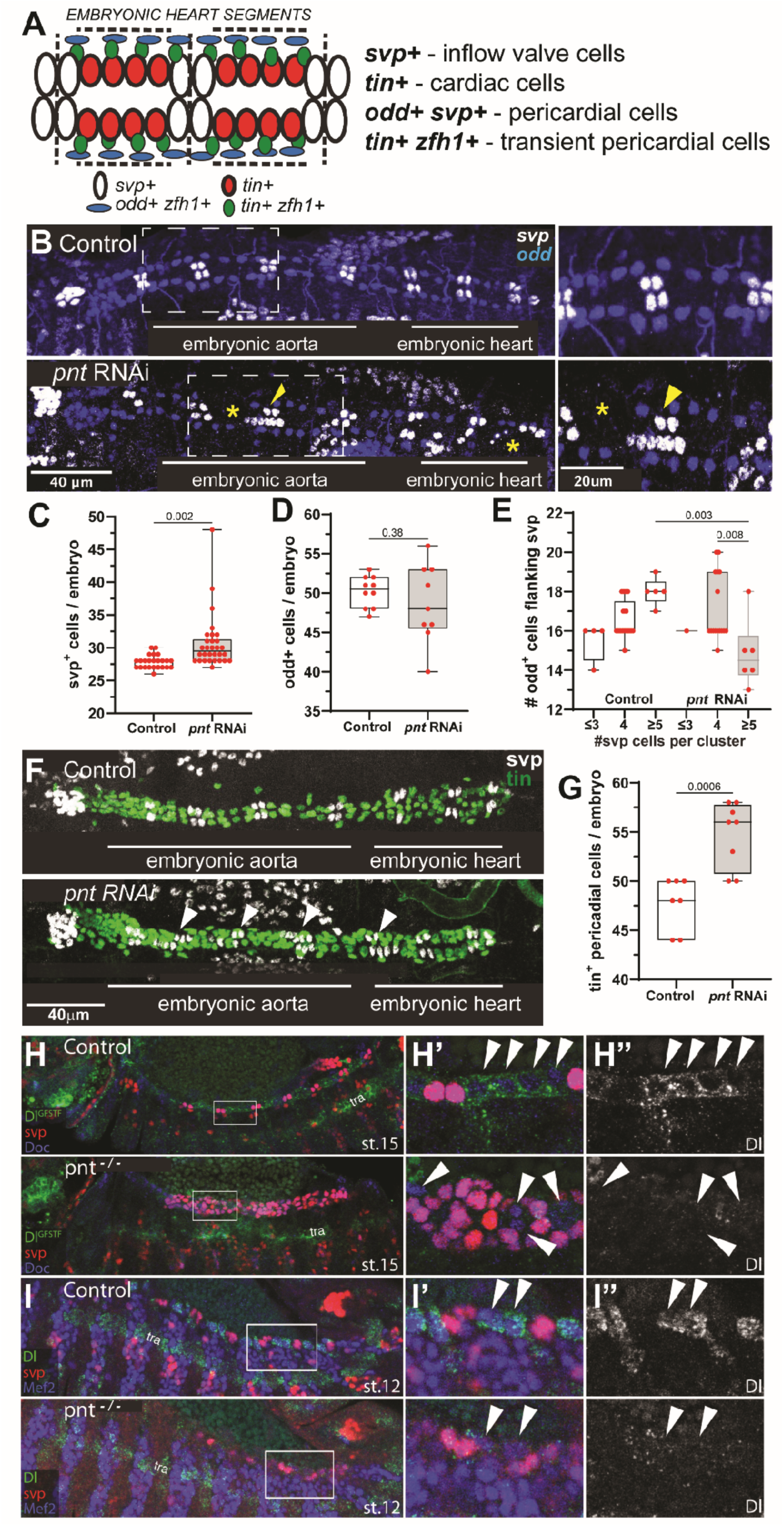
**(A)** Schematic of cardiac and pericardial cell types in the embryonic fly heart. **(B)** Stage 15-17 embryonic fly hearts stained for seven-up (svp) ostial cells (gray) and odd skipped (odd) pericardial cells (blue). Arrowheads mark excess *svp*^+^ cells; * marks missing cells. **(C)** *svp*^+^ cells and **(D)** *odd*^+^ pericardial cells per embryo. **(E)** *odd*^+^/*svp*^+^ ratio in *svp*^+^ clusters. Plots show minimum, first quartile, median, third quartile, and maximum. Significance determined by unpaired Student’s *t*-test. **(F)** Tinman (*tin*^+^) cells (green), quantified in **(G)**. **(H) (Top)** Stage 15 embryos stained for GFP-tagged Delta (Dl), LacZ-tagged *svp*, and Dorsocross (Doc). **(H’ and H”)** Enlarged boxed region showing Dl expression (arrowheads) between *svp*^+^ Doc^+^ inflow valve cells (magenta). **(Bottom)** Stage 15 *pnt* mutant embryos; no Dl expression in *svp* negative cells (arrowheads). **(I) (Top)** Stage 12 *svp-lacZ*/+ embryos with HCR for *Dl* RNA and antibody staining for *svp*-LacZ and Mef2. Arrowheads identify myocardial precursors. **(Bottom)** *svp-lacZ pnt*^Δ88^*/pnt*^Δ88^ mutants, *Dl* is absent from cardioblasts but unchanged in ventral tracheal (tra) precursors.

Immunohistochemistry for *svp^−^ mef2*^+^ cells, which marks working cardioblasts that are normally present as eight cells per segment, showed total *svp^−^ mef2*^+^ cell number was unchanged between controls and *pnt* KD (**Supplemental Figure 2A**). However, among segments with altered *svp^−^ Mef2^+^* cells, controls showed fewer cells, whereas about half of *pnt*-RNAi segments showed increased numbers (**Supplemental Figure 2B**). Total pericardial cell number, marked using *zfh1* and *oddskipped* (*odd*), did not differ significantly between groups (**Figure 5B,D**; **Supplemental Figure 2C-E**), but *odd*^+^ cells were often missing around expanded *svp*^+^ clusters. Stratifying segments by adjacent *svp*^+^ cluster size (≤3, 4, or ≥5 cells) showed that in controls, flanking *odd*^+^ cell number increased with *svp*^+^ number, whereas *pnt* KD showed the opposite trend (**Figure 5E**). By contrast, total *tin*^+^ *zfh1*^+^ pericardial cells were significantly increased in *pnt* KD embryos (**Figure 5F,G**). These findings indicate that mesodermal *pnt* knockdown redirects cardiac cell fate in specific subsets, increasing *svp*^+^ and *svp*^−^ cardioblasts and *tin*^+^ pericardial cells while reducing *odd*^+^ pericardial cells. A second RNAi line driven by *Twist*-GAL4 produced similar but slightly milder effects on *svp*/*Mef2* cell ratios, indicating the phenotype was not due to genetic background (**Supplemental Figure 2F-H**).

Delta-Notch signaling strongly influences cardiomyogenic and pericardial cell numbers in the developing *Drosophila* heart ^24,25^. Because the Notch ligand Delta (Dl) is enriched in precursors of *tin*^+^ cardioblasts ^26^, we tested whether Pnt regulates *Delta*. In controls, Delta was highly expressed in *svp*^−^ cardioblasts, and HCR showed that *Delta* transcription was restricted to precursors of Tin^+^Svp^−^ cardioblasts before their final embryonic division (**Figure 5H,I**). In all *pnt* null mutants, cardioblast *Delta* expression was markedly reduced (**Figure 5H,I**; 30 embryos/genotype). This loss was not explained by conversion of generic Svp^−^ to ostial Svp^+^ cells, because some Svp^−^ generic cardioblasts remained but still lacked normal Dl expression. The effect was tissue-specific, as tracheal precursors retained normal Delta expression. These findings identify Delta-Notch signaling as a major downstream mediator of Pnt function in the cardiogenic mesoderm.

### Ets/pnt KD affects adult fly heart structure and function

We assessed how these embryonic cell-fate changes affect the adult heart using Semi-automatic Optical Heartbeat Analysis (SOHA) ^15^. Mesodermal *pnt* knockdown during early development reduced End Diastolic Diameter (EDD), Fractional Shortening (FS), and cardiac output (**Figure 6A-D),** without altering Heart Period (HP), diastolic interval (DI), systolic interval (SI), or arrhythmicity (**Supplemental Figure 3A-D; Supplemental Table 4)**.

**Figure 6.**
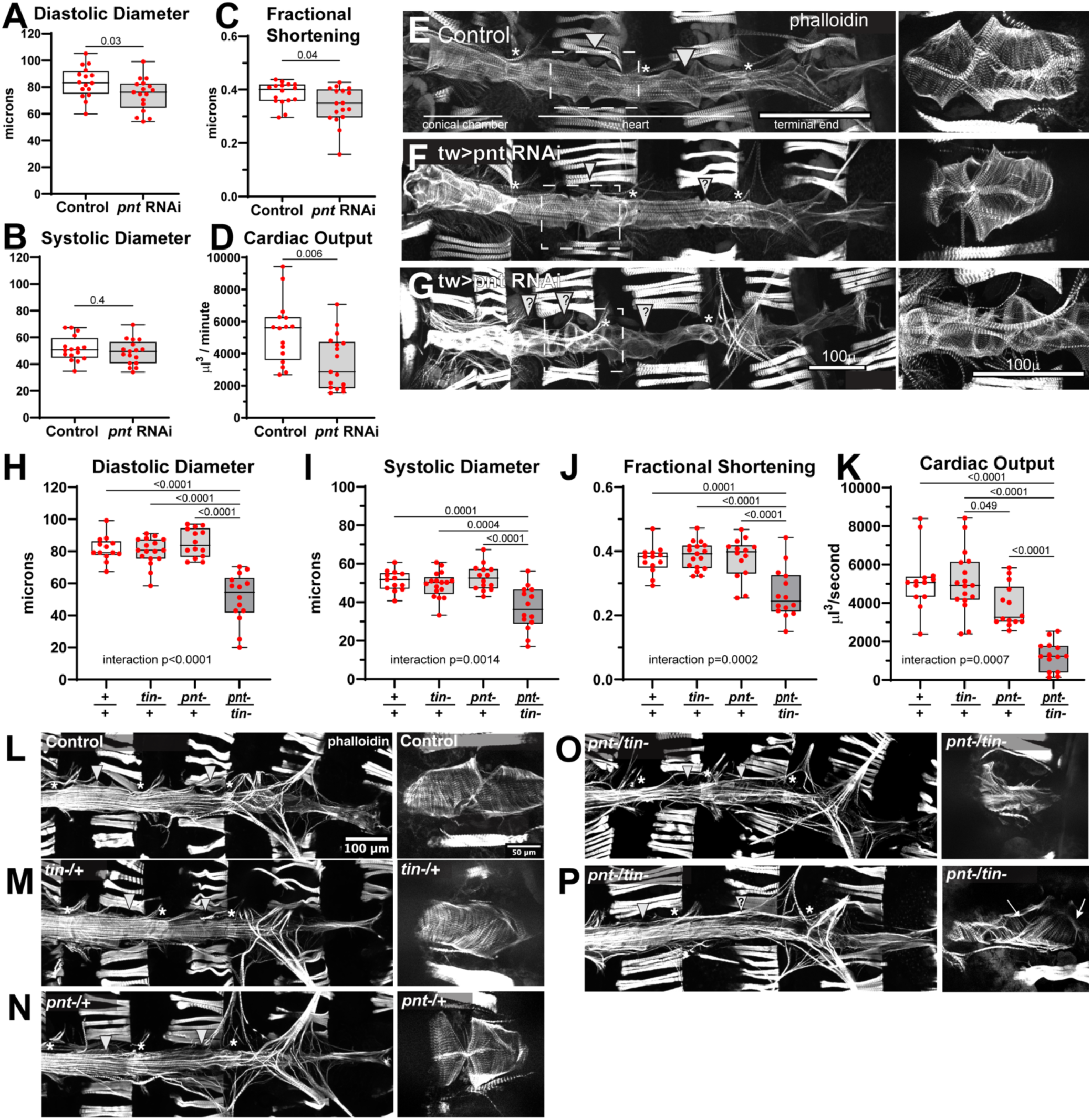
**(A)** End-diastolic diameter, **(B)** end-systolic diameter, **(C)** fractional shortening, and **(D)** cardiac output in control and *pnt* KD (twist-Gal4>UAS-*pnt*RNAi) fly hearts. Plots show minimum, first quartile, median, third quartile, and maximum. Significance by unpaired Student’s *t*-test. **I** F-actin (phalloidin) staining. Controls show well-formed valves (*) and ostia (arrowheads). Mesodermal *pnt* KD caused **(F)** mild and **(G)** severe adult heart defects, with valves and ostia difficult to identify. **(H-K)** Ets/*pnt* interacts with NKX2.5/*tin* to affect **(H)** diastolic diameter, **(I)** systolic diameter, **(J)** fractional shortening, and **(K)** cardiac output in adult hearts. Plots show individual data points with minimum, first quartile, median, third quartile, and maximum. Significance determined by two-way ANOVA with Tukey’s post hoc test. **(L-P)** F-actin (phalloidin) staining of hearts analyzed in **(H-K)**. **(L)** Control hearts and **(M,N)** single heterozygotes; left, full hearts; right, one cardiac chamber. **(O,P)** Double heterozygotes show disrupted circumferential fibers (arrows), inflow tracts (arrowheads), and intracardiac valves (*).

F-actin staining showed that control adult fly hearts had identifiable chambers with circumferentially arranges myofibrils, internal valves, and *svp*^+^ ostial cells (**Figure 6E**). In *pnt* KD hearts a few appeared near normal (**Figure 6F**) but overall the anterior conical chamber was narrower, the posterior end was dilated, and ostia and valve structures were often malformed or difficult to identify. Myofibrils were also highly disorganized (**Figure 6G**). Together, these results indicate that early mesodermal *pnt* knockdown impairs adult heart structure and function, consistent with previous findings in systemic *pnt* mutants ^20^.

### *Ets/pnt* interacts with the cardiogenic transcription factor *tinman/Nkx2.5* in cardiac development

Despite sharing an underlying genetic defect, the cardiac disease phenotypes in humans and in model systems are variable, likely influenced by variable genetic backgrounds. We used the *Drosophila* system to identify genes that interact with *pnt*. To test for genetic interactions, we examined whether combining heterozygous mutations of *pnt* and *tin/Nkx2.5* would lead to worsening of phenotypes compared to controls or single heterozygote mutant phenotypes. Compared to controls, heterozygous *pnt* and *tin* mutants showed no changes in End Diastolic Diameter (EDD), End Systolic Diameter (ESD), FS or CO (**Figure 6H-K**). However, when *pnt* and *tin* mutations were combined as trans heterozygotes, there was a significant decrease in EDD and ESD resulting in significantly reduced FS and CO. In contrast, the presence of a heterozygous *pnt* mutation produced a longer heart period due to longer DIs, but there was no additional effect on HP or DI in the combined *pnt* and *tin* trans heterozygote mutants (**Supplemental Figure 3E-G; Supplemental Table 4**).

Staining with phalloidin made apparent the structural anomalies in response to *pnt* - *tinman* interactions. Hearts from control flies (**Figure 6L**) and both *tin* (EC40/+, **Figure 6M**) and *pnt* (*pntD88/+*, **Figure 6N**) single heterozygotes exhibited similar cardiac structures, including regularly organized circumferential myofibrils within the myocardial cells and easily identifiable ostia (**Figure 6L-N, right**). However, in *pnt* and *tin* trans heterozygote mutants, heart valves and ostial structures were identifiable but clearly malformed (**Figure 6O,P**). F-Actin staining revealed highly disorganized and no longer circumferential myofibrils, with gaps between myofilaments (**Figure 6O,P, right**). These data suggest that moderate reductions in *pnt* and *tin* expression produced by single heterozygous mutations do not significantly alter heart function and structure in adults. However, the combination of these two mutations resulted in significant changes in adult heart function and structure and points to a role for *pnt* within the cardiogenic network driving heart cell identity and heart morphogenesis.

### Characterization of the heart of an infant with Jacobsen syndrome and HLHS

We obtained the heart from a newborn female infant (diagnosed prenatally with a 14Mb terminal deletion in 11q and HLHS) in which comfort care was provided prior to her passing away. A postnatal echocardiogram confirmed a severely hypoplastic, slit-like left ventricle with mitral and aortic atresia (**Figure 1B-E**). Pathologic analysis confirmed the diagnosis (AA/MA, slit-like LV). We also obtained cardiac tissue from an age-matched patient who died from non-cardiac causes. We used immunohistochemistry to characterize and quantify cellular and structural differences in these tissue samples. We stained for the Z band-associated protein α-actinin and DAPI to assess cardiac myocyte structure and organization. Using a blinded protocol, we examined the myofibrillar structure in both right and left ventricles (9 - 22 sections per chamber) from the newborn JS patient with HLHS and the age-matched control heart (**Figure 7A,B**). This qualitative analysis identified more control ventricle slices with regularly arranged Z-bands and identifiable myofibrils (“organized” in **Supplemental Table 3**). This contrasted with heart tissue from the JS patient where it was often difficult to identify sections with distinct Z-banding patterns and myofibrillar structures. Overall, most sections from both right and left JS ventricles showed increased disorganization compared to the control heart (**Figure 7A-D, Supplemental Table 3**). In addition, we noted an increased number of nuclei in patient samples (compare **Figure 7A,B v. C,D**).

**Figure 7.**
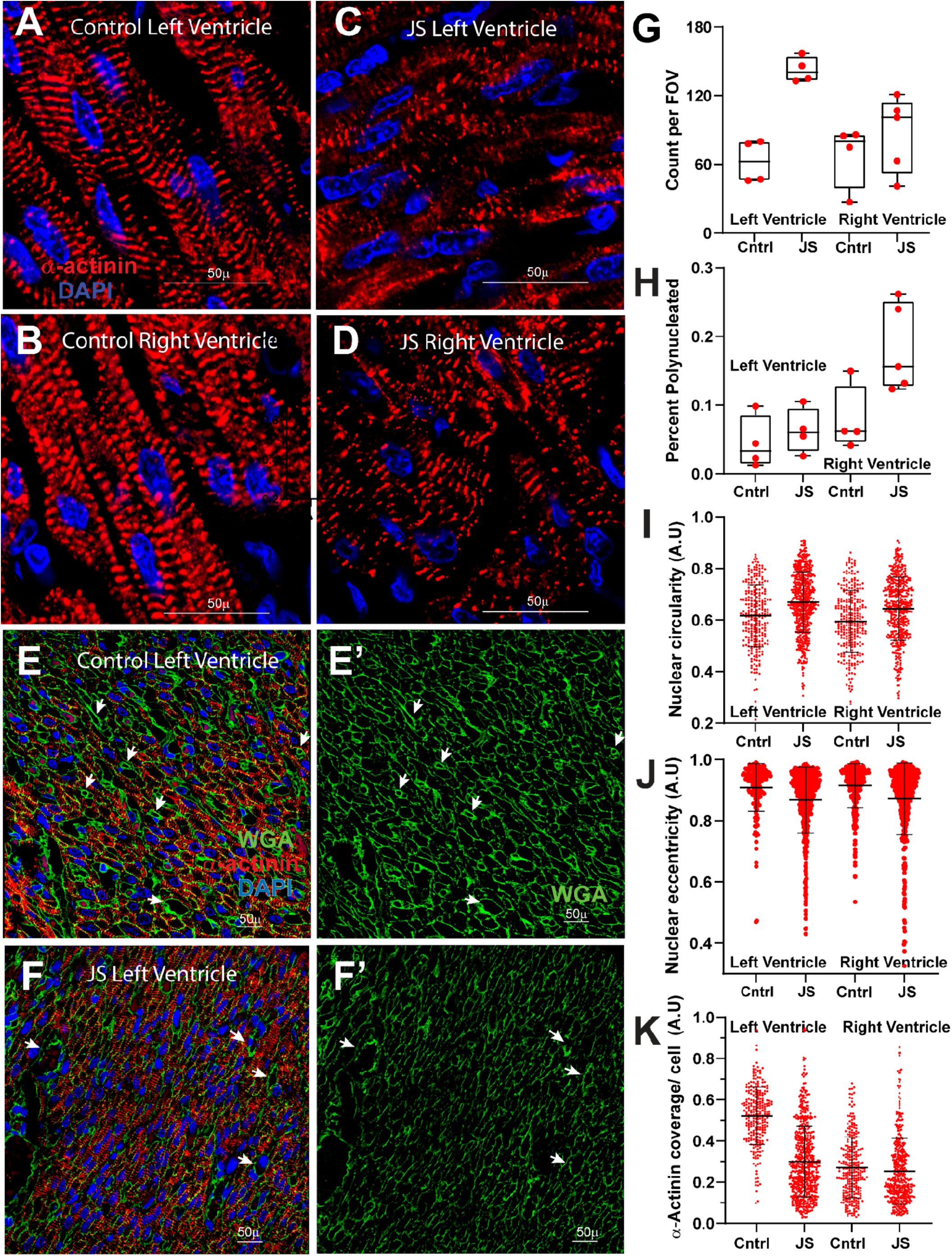
**(A-D)** Ventricular sections from newborn infants stained for α-actinin (red, Z-band) and DAPI (blue). **(A,B)** Left and right ventricles from the Control without congenital heart disease. **(C,D)** Left and right ventricles from an infant with JS and HLHS. **(E-F’)** Images from **(E)** a control heart and **(F)** a newborn JS heart stained for WGA (green), α-actinin (red), and DAPI (blue). Arrows mark membranes around α-actinin-negative structures, likely microvessels. Quantification of myocardial **(G)** cell number, **(H)** percent with polyploid nuclei, **(I)** nuclear circularity, and **(J)** eccentricity from tissue slices located adjacent to the endocardium. **(K)** α-actinin coverage per myocardial cell All plots show mean ± S.D.

To further characterize individual cells, we stained a second set of tissue slices with wheat germ agglutinin (WGA) to visualize cell membranes as well as *α*-actinin and DAPI (**Figure 7 E,F; Supplemental Figure 4).** Qualitatively it appeared that the cell density was increased in the JS LV, but due to the non-continuous nature of the membrane staining it was not possible to accurately identify all individual myocardial cells using automated methods. We therefore used ImageJ ^27^to quantify the total amount of WGA-stained cell membranes in ROIs placed over regions containing primarily a-actinin stained cells as one method to estimate cell numbers (**Supplemental Figure 5A-E**). ROIs from the JS ventricles tended to contain more membrane structures than ROIs from the Control LV (**Supplemental Figure 5F**) suggesting a higher cell density.

To confirm this observation, we used a Python-based custom script (https://github.com/Matthias-Blanc/VAIRONS) and CellPose ^28^ to identify single myocardial cells based on WGA stained (membrane enclosed) structures that contained a-actinin (sarcomeric) staining. There was a trend toward increased number of myocardial cells in the sections from both the left and right ventricle of the JS patient’s heart compared to Control (**Figure 7G**) and nuclei in the JS patient’s myocardial cells tended to be rounder and less eccentric compared to Control (**Figure 7 I,J**). Myocardial nuclei normally become more elliptical (less circular) and located more toward the cell periphery (more eccentric) as myocardial cells mature ^29,30^. We also detected more polynucleated myocardial cells in the JS ventricles (**Figure 7H**) suggesting a premature exit from the proliferative phase ^31^. We also assessed sarcomere structure in these samples by quantifying α-actinin staining, and overall α-actinin cell coverage in myocardial cells tended to be lower in the JS ventricles compared to control (**Figure 7K**), suggesting defects in sarcomere structural development and function.

We observed membrane enclosed structures that did not contain α-actinin in these samples (**Figure 7E,F**, arrows). These structures were easier to identify and appeared more numerous in Control ventricular slices than in the JS patient. We hypothesized that these structures represented the microvasculature within the cardiac tissue and that vasculature was reduced in the JS heart. To test this observation, we immunostained slices for CD31 (PCAM-1), which identifies vasculature endothelium, platelets, megakaryocytes and immune cells. We observed numerous vascular structures in the tissue from control ventricles and very few blood cells (**Supplemental Figure 6A,B**). In the JS ventricles, it was difficult to identify vascular endothelium and most of the staining appeared to be blood cells (**Supplemental Figure 6C,D**). Quantification of the number of vessel structures present in each tissue slice confirmed a relatively reduced number of microvessels in the JS ventricles (**Supplemental Figure 6E**). Further, the abundant blood cells in the JS tissue slices were often seen aggregating together in what appeared to be vessels that lacked stained endothelium (**Supplemental Figure 6C,D**). These findings suggest that the ventricles in the JS patient contain fewer microvessels and with reduced vascular endothelial tissue in the vessels that were present.

## Discussion

We previously identified the ETS1 gene as a candidate gene for causing congenital heart defects in Jacobsen syndrome (JS, OMIM #147791). In the present study, we further investigated the role of the ETS1 transcription factor during early cardiac development using multiple animal model systems. Across frog, zebrafish, and Drosophila models, loss of ETS1 or its orthologs produced a consistent phenotype characterized by hypoplastic cardiac chambers and impaired cardiac function. The defects in cardiac tissue organization include abnormal myofibrillar organization in both human and fly hearts, endocardial misalignment in both frog and zebrafish, and loss of the trabecular myocardium in fish and frog. In addition, our studies on human heart tissue from a patient with JS and HLHS implicate ETS factors for normal coronary vascular endothelial development, paralleling our previous studies in mice ^6^. These data support a model in which ETS1 deficiency causes primary defects in cardiac endothelial development early in cardiogenesis, leading directly to impaired ventricular growth.

### Loss of ETS factors disrupts cardiomyocyte development and causes ventricular hypoplasia

Knockdown or knockout of ETS factors reduced cardiac size in all four model systems. In frog, zebrafish, and mouse, ventricular hypoplasia was accompanied by loss of trabecular myocardium, resembling the slit-like left ventricle seen in the aortic atresia/mitral atresia (AA/MA) subtype of HLHS. Several mechanisms may contribute to this reduction in ventricular size. In zebrafish, ventricular hypoplasia was associated with fewer total cardiac cells, suggesting impaired proliferation. This reduction was more pronounced in endocardial cells than in cardiomyocytes, pointing to an early defect in cardiac cell fate specification. This interpretation is supported by our *Drosophila* studies, in which mesodermal knockdown of the ETS ortholog *pointed* altered cardioblast identity during early heart specification (Figure 6). Together, these findings suggest that early defects in cell fate determination contribute to the later loss of trabecular myocardium.

Our current fly studies, together with our previous mouse work ^6^ implicate Notch signaling in early cell fate specification. In frog, reduced *kdr* (VEGF2) staining further suggests that ETS loss disrupts key pathways involved in early cardiac endothelial specification and proliferation. Notably, transplantation of wild-type heart field tissue into ETS1-deficient frog embryos restored ventricular morphogenesis and normalized endocardial organization, indicating that ETS1 functions early within cardiac mesoderm to regulate tissue migration and development. These results support the hypothesis that defective endocardial development and impaired endocardial-myocardial signaling are major contributors to the HLHS ventricular phenotype, consistent with previous studies by Miao et al ^32^.

Another contributing mechanism may involve premature cardiomyocyte cell-cycle exit. Ventricular tissue from a patient with JS/HLHS showed a dramatic increase in nuclei in both ventricles compared to age-matched control tissue, which along with reduced nuclear size and decreased sarcomeric structure, is consistent with altered cardiomyocyte maturation. This interpretation aligns with our earlier mouse studies showing that endothelial-specific deletion of *ETS1* caused premature maturation and reduced proliferation in a subset of cardiomyocytes within the compact myocardium^7^.

### Endocardial development is essential for ventricular morphogenesis

Our mouse studies provide direct evidence that genetic ablation of the endocardium causes ventricular hypoplasia, paralleling the ETS-deficient phenotypes observed in frog and fish, implicating a requirement for endocardium in ventricular development. Although the endocardium is already known to regulate trabeculation through multiple signaling pathways, direct ablation of this lineage using the *Nfatc1Cre*;DTA system links the physical absence of endocardial cells to ventricular hypoplasia and loss of the trabecular myocardium. This model therefore offers a useful *in vivo* framework for studying the AA/MA/slit-like LV subtype of HLHS.

In mice, endothelial deletion of ETS1 led to ventricular non-compaction and decreased levels of cardiac factors that promote compact zone cardiac myocyte proliferation {Wang, 2026 #32}. Future studies will determine whether these same growth factors are deficient in the developing HLHS left ventricle, and what the species-specific factors are that determine ventricular non-compaction vs. hypoplasia. The species-specific difference in ventricular phenotypes suggests that loss of *Ets1* alone may be insufficient in mice, and possibly in humans, to fully disrupt early ventricular development and trabeculation. We hypothesize that functional redundancy among ETS family members, including *Ets2* and *Etv2*, may partially compensate for the loss of *ETS1* during mammalian cardiac development ^33^. This is supported by our observation in zebrafish that the combined KO of Ets1,2 and Etv2 caused a more severe phenotype than did KO of Ets1 alone.

### Human JS/HLHS heart tissue reveals maturation defects and reduced coronary microvasculature

Although *ETS1* mutations have been linked to Jacobsen syndrome and HLHS, detailed histological analysis of patient tissue provides important new pathological insights. Our observations of increased myocardial cell numbers combined with decreased a-actinin area/cell and rounder nuclei suggest that the myocardial cells in the JS left ventricle were smaller and more densely packed with reduced development of sarcomeric structures, indicative of impaired myocardial maturation. Consistent with that observation, the increase in polyploid cells was more striking in the right ventricle, suggestive of a differential effect on cardiac myocyte proliferation between the two ventricles. Our findings of reduced coronary microvasculature along with reduced vascular endothelium are consistent with our previous mouse studies demonstrating that ETS1 is required in both the endocardium and coronary vascular endothelium for normal cardiomyocyte proliferation and compact-zone growth ^7^. A dual role for these endothelial lineages has also been proposed in earlier studies of human HLHS. Miao et al. ^32^ showed that endocardial cells derived from HLHS patient iPSCs have impaired ETS1-dependent function, and Yu et al. ^34^ identified abnormalities in the coronary vascular endothelium and Notch signaling in HLHS patients with Kabuki syndrome. Collectively, these studies support a model in which defective endocardial and/or coronary vascular endothelial signaling disrupts ventricular morphogenesis early in development.

### ETS factors interact with additional cardiac developmental pathways

Across model systems, our results indicate that ventricular hypoplasia arises from defects early in heart development and that ETS factors are central components of this process. At the same time, phenotypic variability across embryos and species suggests that other pathways modify the effects of ETS1 deficiency. In *Drosophila*, we identified a novel interaction between *pnt* and *tinman*, the ortholog of *Nkx2.5* ^35^. Although both genes were already known to contribute to cardiogenesis, their synergistic interaction has not been previously characterized. Trans-heterozygous mutants showed combined defects in adult heart structure and function, revealing a conserved regulatory network linking ETS signaling with *Nkx2.5*/*tinman* pathways. In addition, loss of *pnt*/ETS disrupted Notch signaling, suggesting further integration with this developmental pathway. Reduced Notch activity has been reported previously in HLHS patient-derived endocardial iPSCs ^32^and more recently in ETS1 knockout mice ^6^. These findings suggest that ETS factors function within a highly conserved developmental network integrating Nkx2.5/tinman and Notch signaling during cardiac morphogenesis.

### Implications for the pathogenesis of HLHS

Our findings highlight the importance of developmental timing in ventricular hypoplasia, especially in the AA/MA subtype of HLHS. Across model organisms, ETS1 deficiency caused early defects in cardiac development that led to ventricular hypoplasia. This differs from late mitral inflow obstruction models in chick ^36^ and mouse ^37^ models, in which chamber obliteration coincides with myocardial overgrowth (“no flow/overgrow”). Instead, our data support a model in which the hypoplastic ventricle arises from impaired outward ventricular growth (“ballooning”) early in development due to primary defects in cardiac vascular endothelial and endocardial function. These defects likely impair trabecular formation and limit ventricular expansion, which may help explain why fetal relief of aortic stenosis often does not prevent progression to HLH^38^. Although ventricular endothelial dysfunction appears primary, abnormal valve development may also contribute. In fly, both inflow tract and inter-chamber valve development was impaired (**Figure 6**) and in zebrafish both the atrioventricular canal and outflow tract development were affected (**Table 3**). Because ETS1 is expressed in developing mitral valve endocardium in mice, early mitral inflow obstruction may contribute to the phenotype. We therefore propose a three-hit model for HLHS in Jacobsen syndrome:

1. Early mitral obstruction caused by abnormal mitral valve development due to impaired valvar endothelium.
2. Impaired ventricular endothelial function affecting both endocardial and coronary vascular endothelial signaling/growth factor secretion, leading to decreased ventricular growth due to decreased cardiac myocyte proliferation.
3. Impaired cardiac myocyte sarcomeric structure and alignment, preventing normal growth and development of the trabecular myocardium and ventricle.

Together, these findings support a conserved role for ETS factors in early ventricular morphogenesis and help bridge the gap between animal models of ETS1 deficiency and human disease pathology.

## Supporting information

Supplemental Movie 1 Frog Control_FLFM

Supplemental Movie 2 Frog Ets MO_FLFM

Supplemental Movie 3 Ets1 KO

Supplemental Movie 4 Fly Ets1 2 Etv2

## Data Availability

All data produced in the present work are contained in the manuscript.

## Supplementary material

Supplementary material is available online.

## Acknowledgements

We want to acknowledge the UCSD School of Medicine Microscopy Core (Grant: NS047101) for invaluable technical support and Dr. Wenhao Liu for invaluable technical support and for assisting in the FLFM image acquisition and analysis.

