## Supplemental Material and Methods for "Conserved roles of cardiac endothelial ETS factors in ventricular development and disease"

#### Limitations:

A limitation of the fly model is that it does not have a ventricle per se, although the conical chamber has been proposed to serve a ventricle-like function. However, the conservation of heart development genetics (e.g. Nkx2.5 was first identified in the fly [1] combined with the low genetic redundancy has allowed us to study the role of Ets/pnt and genetic interactions in a relatively straightforward manner.

While changes in cell size and number are important features of ventricular hypoplasia, we were unable to quantitatively measure that in frog embryos due to relatively bigger embryo sizes and tissue opacity. We can only speculate based on our results from zebrafish embryos that there may be a decrease in the number of cardiac cells in the significantly smaller heart of mutant embryos.

Our human data are based on only two age-matched human hearts, but due to very limited patient material we nevertheless decided to perform a detailed assessment of the myocardial organization as an initial analysis. We acknowledge that this represents an N of one for the JS HLHS patient and control. For this reason, we have not attempted to perform any statistics and have only indicated apparent trends in the data. Nevertheless, these data inform and are consistent with numerous studies demonstrating abnormal coronary vasculature in HLHS hearts of unknown etiology [2] and studies in chick demonstrating a critical role for ETS factors in coronary vascular development [3]

While our models do not recapitulate the human disease (i.e., given that frogs and zebrafish only have one ventricle), our studies demonstrate that there are significant similarities in phenotypes as well as conservation of genetic function. Importantly, it is the combination of information from these models that will permit the identification of the genetic interactions and cellular mechanisms that impact HLHS in humans.

#### Supplemental Material and Methods:

<u>Frog methods:</u>

*Frog embryo manipulations, morpholino oligomer, and RNA preparation:* wild type or transgenic embryos of *Xenopus laevis* were obtained and staged per the Normal Table of *Xenopus laevis* [4]. The embryos were microinjected with capped RNA or morpholino oligomer (MO) during early cleavage stages. Ets1-MO (5'- taa ggt cta gtg cag ctt tca tgg c -3') targeting the translation start site of Ets1 transcripts has been described previously by Nie and Bronner, 2015 [5] and was injected at 10ng into each side of the 4-cell stage embryo (20ng total). The specificity of the morpholino has been verified previously [5]. The latest stage for our analysis was ~stage 46, well before the time when morpholinos have lost their effect due to dilution. RNAs of nuclear GFP (H2b-EGFP) were synthesized with linearized templates using SP6 polymerase (Ambion mMessage mMachine Kit) and co-injected into embryos at 0.1-0.2ng per embryo when needed. To obtain transgenic embryos, sperms from myl3:GFP or kdr:GFP transgenic animals (heterozygous) were purchased from NXR (National Xenopus Resource) and used to fertilize wild type oocytes. Since only about half of the resulting embryos carry the transgene, embryos were sorted at tailbud stages. Myl3:GFP labels cardiomyocytes and skeletal muscles, while kdr:GFP labels endocardial and endothelial cells [6,7]. All experimental procedures were performed according to USDA Animal Welfare Act Regulations and have been approved by the Institutional Animal Care and Use Committee, in compliance with Public Health Service Policy.

*Heart field transplantation:* Donor embryos injected with H2b-EGFP were sorted under fluorescent microscope for good signal in the heart field. These embryos and stage matched host embryos injected with Ets1-MO were transferred into MBSH (high salt buffer to support healing) on agarose beds. 1% MS222 was added in the medium to anesthetize the embryos. Heart field tissue was isolated from donor embryos and grafted into host embryos with the heart field tissue removed [8]. The embryos were allowed to heal for about an hour before they were transferred back into 0.1xMMR without MS222.

*Immunohistochemistry and in situ hybridization:* Immunohistochemistry analysis of wild type, mutant, and transplanted embryos were performed as previously described [9]. Anti-Trypomyosin antibody (CH1; DSHB) and anti-MYH1E antibody (MF20; DSHB) were used at ~1-5μg/ml, and fluorescent secondary antibodies were used at 1:500 dilutions. DAPI staining was performed after antibody staining when needed. Hybridization Chain Reaction amplified *in situ* hybridization (HCR *in situ*) was performed by modifying the protocol from LaBonne lab [10]. Briefly, fixed embryos were rehydrated, briefly treated with proteinase K, rinsed, and hybridized with probes sets for *pecam1* (Molecular Instruments) overnight at 37°C. Excess probes were removed by extensive washes and then incubated with DNA hairpins labeled with Alexa 546 (Molecular Instruments) for amplification. Washed embryos were then counterstained with Dapi. Embryos were cleared in BA/BB and mounted between cover glasses and imaged under Nikon W1 Spinning Disk Confocal microscope.

*Fourier light-field microscopy (FLFM), image reconstruction, and analysis:* Wild type or mutant tadpoles receiving H2b-EGFP in cardiogenic mesoderm were raised to stage 46 and mounted with ventral side up on agarose bed in 0.1xMMR plus 0.1%MS222. They were then subject to FLFM imaging [11] for 1 minute. Volumetric reconstruction of the imaging data was performed through a wave-optics-based Richardson-Lucy deconvolution of the elemental images and point-spread function [12]. Heart rate and ventricular chamber size at contraction and relaxation were measured, from which ejection fraction (1 - contraction volume/relaxation volume) and cardiac output (heart rate × (relaxation volume - contraction volume)) were calculated and compared. Student *t-test* was performed to determine statistical significance.

Zebrafish methods*:*

Assays in zebrafish: In-depth quantitative analyses of zebrafish cardiac function and conduction dynamics was performed as described [13]. Larval zebrafish at 24-72 hpf were immobilized in a small volume of low melt agarose (1.5-2%) and submerged in conditioned water. Hearts were imaged *in vivo* at room temperature (20-21$^{o}$C) with direct immersion optics and a digital high-speed camera (Orca Flash, Hamamatsu Photonics). High-speed movies (~150 fps) were analyzed using SOHA [13] to quantify heart period (R-R interval), chamber size, and contractility (fractional area change). Cardiac stroke volume was estimated using 2D chamber size and assuming a prolate ellipsoid chamber shape. Cardiac output was calculated as the stroke volume times heart rate (1/heart period).

*CRISPR in zebrafish*: *ets1, ets2* and *etv2* were all targeted for mutagenesis using CRISPR/Cas9 genome editing to induce indel mutations in candidate genes [14]. We used mixed single guide RNAs (sgRNAs) composed of the targeting sequence that is followed by a PAM sequence that guides the Cas9 nuclease to the desired genomic locus [15].

| gene name | accession number | IDT | gRNA |
|---|---|---|---|
| ets1 | NM_001017558.1 | **Dr.Cas9. ETS1.1.AE** | GGCCUUCUGUCCACUCACGC |
| ets2 | NM_001023580.1 | **Dr.Cas9. ETS2.1.AB** | ACGUCUGGAAGAGCUCUCGC |
| etv2 /etsrp | NM_001037375.1 | **Dr.Cas9. ETV2.1.AA** | UGGUCCGACUACCCCUCACC |

*Zebrafish Immunohistochemistry:* To identify developmental/morphological defects we used *Tg(myl7:EGFP)* as a reporter to evaluate the overall heart chamber shape and H2A-mCherry expression in the nuclei (*Tg(myl7:H2A-mCherry*) [16] to quantify cardiomyocytes from reconstructions of confocal Z stacks. Fish were fixed in 4% paraformaldehyde, stained with anti-GFP and anti-mCherry antibodies (Thermofisher). Anti-myosin antibody was MF-20 (dshb.biology.uiowa.edu/MF-20, Developmental Studies Hybridoma Bank). Hearts were imaged with a confocal microscope (Zeiss LSM-710). Cell counts were made using Imaris image analysis software (v9.8).

<u>Mouse Methods</u>:

*Immunofluorescence:* Mouse embryos were dissected in PBS and fixed overnight at 4 °C in 4% PFA. After fixation, they were sequentially incubated in 5%, 10%, 15%, and 20% sucrose in PBS, embedded in OCT Tissue-Tek (Thermo Fisher Scientific), and sectioned into 8-μm slices using a Leica CM 3050S Cryostat (Leica Microsystems). Sections were blocked with 10% donkey serum in 0.1% PBST (PBS with 0.1% Triton-X 100) at room temperature for 1 hour, followed by overnight incubation at 4 °C with primary antibodies (Rabbit mAb anti-ERG, Abcam, Ab92513; Mouse mAb anti-a-Actinin, Sigma-Aldrich, A7811) diluted in blocking solution. After three washes in 0.1% PBST at room temperature, sections were incubated with secondary antibodies and DAPI (1:1,000) at room temperature for 1 hour. Sections were then washed three more times in 0.1% PBST and mounted in DAKO fluorescence mounting medium (Agilent). Immunofluorescence images were acquired using a Zeiss LSM 880 Airy Scan Confocal Microscope.

<u>Fly methods:</u>

*Drosophila Strains*: Heart-specific control of transcription was achieved by the GAL4-UAS system in which the following *Twist*-GAL4 and *Hand4.*2-GAL4 drivers were used to knockdown genes. *Drosophila* GD and KK RNAi collection lines along with appropriate controls were obtained from the Vienna Drosophila Resource Center (VDRC) [17]. All fly lines are listed in **Supplemental Table.**

*Embryo Collections and Stainin*: Embryos were collected and stained as previously described [18]. Briefly, Flies were placed in cages with removable grape juice agar plates smeared with rehydrated Baker's yeast. Plates were collected after about 17 hours, dechorionated for 3 mins in 5% Bleach and transferred to 1.5 mL Eppendorf tubes containing fixative (2 parts heptane, 1part 1x PBS, 1part 4% formaldehyde). Embryos were fixed for 20 mins on a shaker. The lower fixative layer was removed and replaced with methanol and vortexed for 30 seconds. Then all liquid is removed, replaced with methanol and incubated on a shaker for 1 hour or until embryos are ready for staining. The methanol is then replaced with 1x

PBT and washed for 10 mins and repeated three times. Embryos were incubated with primary antibodies diluted in 1X PBT overnight at 4ºC then washed with 1X PBT over 2 hours. Embryos were then incubated with secondary antibodies diluted in 1X PBT and then washed with 1X PBT over 2 hours. Before mounting using ProLong Gold Mountant with DAPI (Life Technologies), embryos were washed once with 1X PBS. All antibodies used in these studies are listed in Supplementary Table __.

*Heart Function Analysis*: Assessment of *Drosophila* heart function and structure using the Semi-automatic Optical Heartbeat Analysis (SOHA) method as previously described [13]. Briefly, four-day old adult flies were anesthetized with FlyNap (Carolina Biological Supply Co, Burlington, NC) and dissected in oxygenated artificial hemolymph to expose the beating heart within the abdomen. Hearts were placed on an Olympus BX61WI microscope while being filmed through a 10x water immersion lens with a high-speed digital camera (Hamamatsu Photonics C9300 digital camera) using HCI image capture software (Hamamatsu). High-speed movies were analyzed using the Semi-automated Optical Heartbeat Analysis (SOHA) software [18, 33]. Parameters measured include Heart Period (HP), Diastolic Interval (DI), Systolic Interval (SI), Arrhythmicity Index (AI), Diastolic Diameter (DD), Systolic Diameter (SD) and Fractional Shortening (FS), a measure of contractility, is calculated using the following equation FS=(DD-SD)/DD.

*Statistical Analyses*: Raw data was blinded prior to functional analysis. Drosophila cardiac function data was analyzed using the D'Agostino and Pearson omnibus normality test for Gaussian distribution. For data that were distributed normally, statistical significance was determined using a 1-way ANOVA for simple comparisons and 2-way ANOVA for multiple manipulations followed by multiple comparisons post-hoc tests. Data sets that do not show a normal distribution was analyzed using a nonparametric 2-tailed unpaired t-test, Wilcoxon Rank Sum test, or Kruskal-Wallis test followed by Dunn multiple comparisons post-hoc tests. Genetic interactions were identified using two-way ANOVA with appropriate post-hoc tests to further confirm significant "interactions".

*Immunostaining of Adult Drosophila Hearts*: Adult flies were dissected and treated with 10mM EGTA in PBT (PBS + Triton-X-100; 0.03% Triton X-100), for 2 minutes to maintain a relaxed state of the heart. Hearts were then fixed with 4% PFA in PBT for 20 minutes, followed by three 10-minute PBT washes. Hearts were stained with primary antibodies (EC11 *Pericardin*, Developmental Studies Hybridoma Bank, DSHB) and incubated overnight in 4oC. Hearts were then washed with PBT three times for 15 minutes each, followed by incubation with fluorescent secondary antibodies (1:500, Jackson ImmunoResearch Laboratories, Inc.) and Alexa Fluor conjugated phalloidin (1:300, Life Technologies) at 4ºC overnight. Hearts were then washed with PBT three times for 15 minutes each and then once with PBS. Hearts were mounted using ProLong Gold Mountant with DAPI (Life Technologies). Delta RNA was detected via hybridization chain reaction as in Choi et al., 2018 [19] with amplifiers and probes (Dl lot # PRH497) ordered from Molecular. Immunostained preparations were visualized with an Imager.Z1 equipped with an Apotome2 (Carl Zeiss, Jena), Hammamatsu Orca Flash4.0 camera, and ZEN imaging software (Carl Zeiss).

Human Methods:

*Immunofluorescence*: SectionsSlides were baked at 60 °C for 60 min, then deparaffinized by incubation in xylene (2 × 5 min), followed by 100% ethanol (2 × 3 min), 95%, 80%, and 70% ethanol (3 min each), and finally rinsed in dd$H_2O$ for 2 min. For antigen retrieval, slides were placed in citrate buffer (pH 6.0), brought to near-boiling and maintained for 15 min, cooled at room temperature for 30 min, and washed in PBS (3 × 5 min). Sections were permeabilized in 0.1% Triton X-100 in PBS for 15 min at room temperature, then blocked with 10% donkey serum in 0.1% PBST (PBS with 0.1% Triton X-100) for 1 h at room temperature. Primary antibody (Rabbit mAb anti-Sarcomeric Alpha Actinin, Abcam, ab68167) diluted in the blocking solution was applied and incubated overnight at 4 °C, followed by three washes in 0.1% PBST. Sections were then incubated with fluorescent secondary antibodies and DAPI (1:1,000) for 1 h at room temperature, washed again three times in 0.1% PBST, and mounted using DAKO fluorescence mounting medium (Agilent). To validate antibody specificity, an IgG isotype control from the same species was used in place of the primary antibody, and secondary antibody-only controls were included to distinguish true target staining from background. Immunofluorescence images were acquired using a Zeiss LSM 880 Airy Scan Confocal Microscope at 20X magnification.

Identification and quantification of cell membranes from WGA stained images was performed using FIJI ImageJ Particles Analysis function [20].

The qualitative assessment of the myofibrillar and Z band organization was performed blinded on α-actinin stained slices. All images were assigned a code using a random number generator and assessed on a three point scale (1-Parallel myofibrils/regularly spaced Z bands, 2-nonparallel myofibrils /disorganized Z-bands, 3-Mixture of organized and disorganized myofibrils/Z bands) by blinded researchers using two exemplar images per category as a guide. Results from two blinded assessments were averaged.

*Image analysis*: Single-cell quantification was performed using Varions (https://github.com/Matthias-Blanc/VAIRONS), an open-source Python application built on NumPy (v2.5 [21]), SciPy (v1.17[22]), scikit-image (v0.26, [23]), pandas(v2.3) and Cellpose (v4.2, [24]) under Python 3].

*Segmentation and Quantification*: Nuclei, cytoplasm and α-actinin area were segmented using the Cellpose-SAM ("cpsamv2") deep-learning model [23], objects smaller than 200px for actinin, 2000px for nuclei, 3000px for cytoplasm or lying within 5px of the image border were discarded. Each nucleus was automatically matched to its cytoplasm by majority assignment within a 2-px ring around it. Analyses were restricted to cells positive for alpha-actinin. A cell was retained when the marker's binary mask covered the cell by a the area of an average alpha actinin particle. For every analyzed cell, the stained area ($\mu m^2$), the number of stained particles, and the fraction of the compartment area covered by the stain (coverage) were quantified. For each segmented nucleus the following features were quantified and exported: morphology area and perimeter (calibrated to $\mu m^{\wedge 2}$ and µm respectively), circularity, eccentricity, solidity, major and minor axis lengths, and the number of convexity defects. Nuclear alignment was summarized per field as the axial order parameter (the mean resultant length of the doubled nuclear orientation angles), ranging from 0 (random orientation) to 1 (perfect alignment).

*Outlier removal*: Starting from the selected cells, any cell whose robust z-score, |x - median| / (1.4826 x MAD), exceeded [10] on at least one quality-control feature was removed; the quality-control feature set comprised object morphology, within-region intensity (mean and standard deviation) and marker magnitude (stained area, coverage and particle count), and deliberately excluded per-channel mean intensities and texture so that genuinely marker-bright cells were retained. Features with zero median absolute deviation, and missing values, were ignored.

### Supplemental Figures

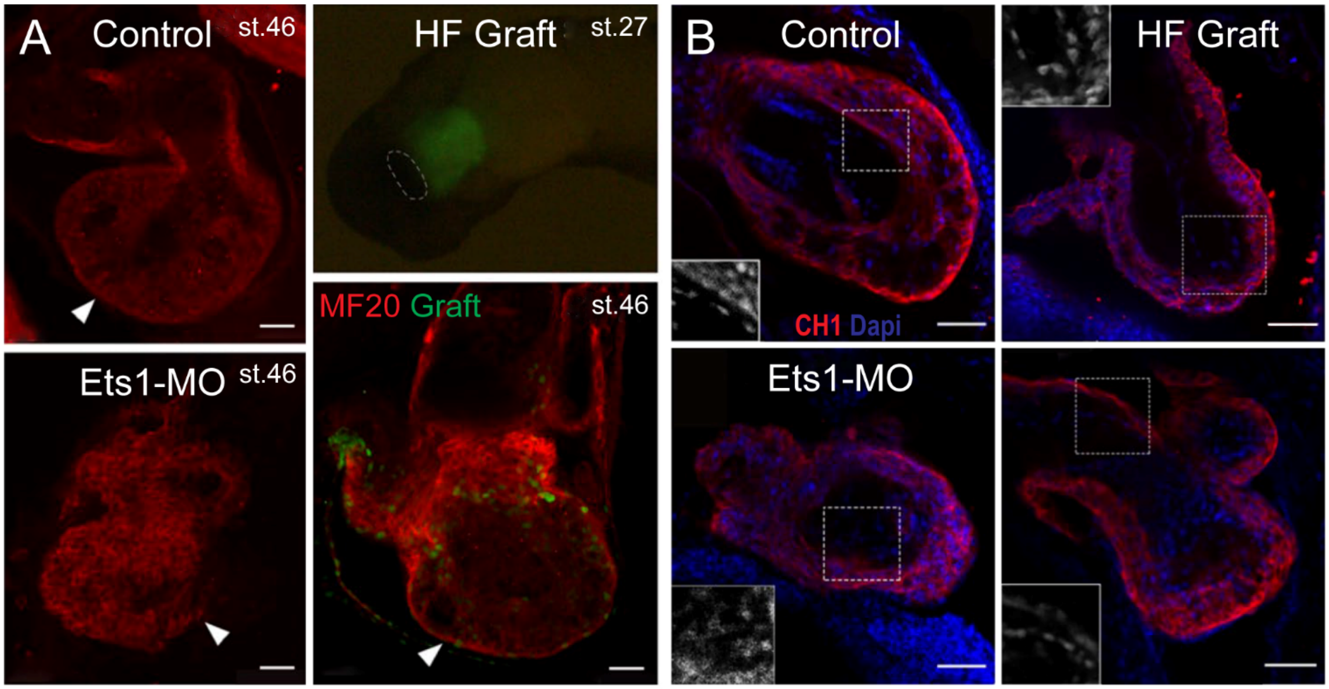


**Supplemental Figure 1:** Heart field transplantation into Ets1 morphant embryos at stage 24 restores normal frog heart development. **(A)** Top left, control heart at stage 46. Top right, wild-type heart field tissue expressing GFP (green). Cardiomyocytes were labeled with an antibody against myosin heavy chain (red). Bottom left, defective ventricular development in Ets1-MO-injected tadpoles. Bottom right, normal ventricular development in Ets1-MO-injected tadpoles transplanted with wild-type GFP-expressing heart field tissue. **(B)** Endocardial cell alignment was also rescued. Hearts at stage 41 were labeled for cardiomyocytes (tropomyosin, CH1, red) and nuclei (DAPI, blue). Inset shows DAPI staining from the boxed areas. Scale bars, 50 µm.

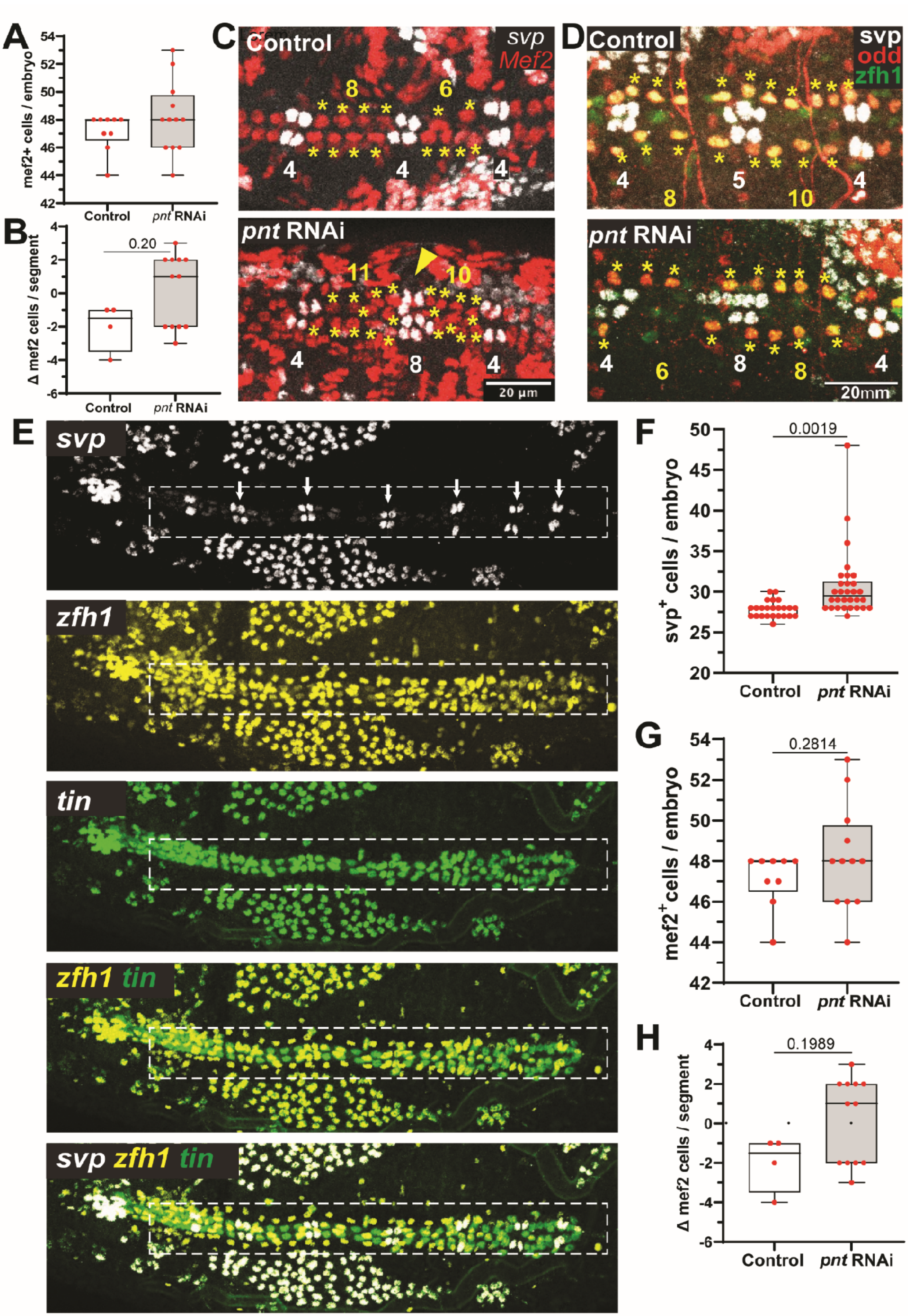
A
mef2+ cells / embryo
Control
pnt RNAi
B
Δ mef2 cells / segment
0.20
C
Control
svp
Mef2
pnt RNAi
20 µm
D
Control
svp
odd
zfh1
pnt RNAi
20mm
E
svp
zfh1
tin
zfh1 tin
svp zfh1 tin
F
svp+ cells / embryo
0.0019
G
mef2+cells / embryo
0.2814
H
Δ mef2 cells / segment
0.1989

**Supplemental Figure 2: (A)** Control hearts usually contain eight *Mef2*$^+$ cardiac cells per segment, with no difference in total *Mef2*$^+$ cell number between control and *pnt*-RNAi embryos. **(B)** In segments with altered *Mef2*$^+$ counts, controls usually had fewer cells, whereas *pnt*-RNAi embryos often had more. **(C)** *svp*$^+$ and *Mef2*$^+$ staining in control and *pnt*-RNAi embryos. * marks *Mef2*$^+$ cells; yellow numbers indicate *Mef2*$^+$ cells; white numbers indicate *svp*$^+$ cells. **(D)** Posterior embryonic hearts from control and Twist-GAL4>*pnt* KD embryos. * marks *odd*$^+$ cells; yellow numbers indicate *odd*$^+$ cells; white numbers indicate *svp*$^+$ cells. **(E)** Paired *svp*$^+$ cells are ostial cells (inflow tracts, arrows); *zfh1*$^+$ cells are pericardial cells; *tin*$^+$ cells lacking *zfh1* are cardioblasts. **(F)** Twist-GAL4 knockdown using a second *pnt*-RNAi line (VDRC 7171) also increased overall *svp*$^+$ cell counts. **(G)** Total *Mef2*$^+$ cell number again did not differ between control and *pnt*-RNAi embryos. **(H)** However, in segments with altered *Mef2*$^+$ counts, many *pnt*-RNAi embryos had increased cell numbers, similar to *pnt*-RNAi line 105390 in **(A,B)**. Plots show minimum, first quartile, median, third quartile, and maximum. Significance by unpaired Student's *t*-test.

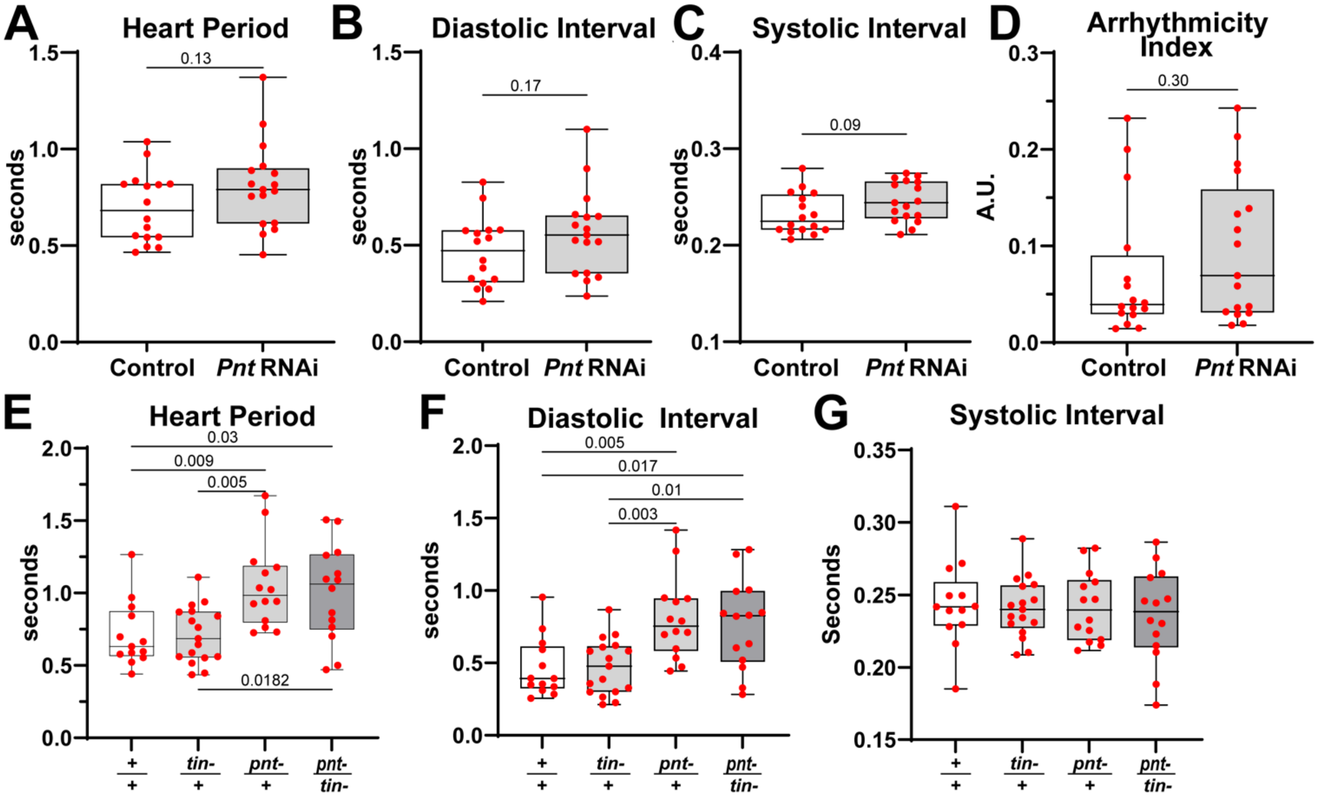


**Supplemental Figure 3:** Knockdown of *ETS1/pnt*. All plots show individual data points with minimum, first quartile, median, third quartile, and maximum. **(A-G)** Mesodermal KD of *ETS1/pnt* did not alter temporal parameters including **(A)** heart period, **(B)** diastolic or **(C)** systolic interval, or **(D)** arrhythmia. A-H) Significance by unpaired Student's *t*-test. **(E-G)** Heart period, diastolic interval, and systolic interval in hearts from wild type (+/+), *Nkx2.5/ tinman* heterozygotes (*tin-*/+), *ETS1/pnt* heterozygotes (*pnt-*/+), and double heterozygotes (*pnt-/tin*). (I-K) Significance by two-way ANOVA with Tukey's multiple-comparisons post hoc test.

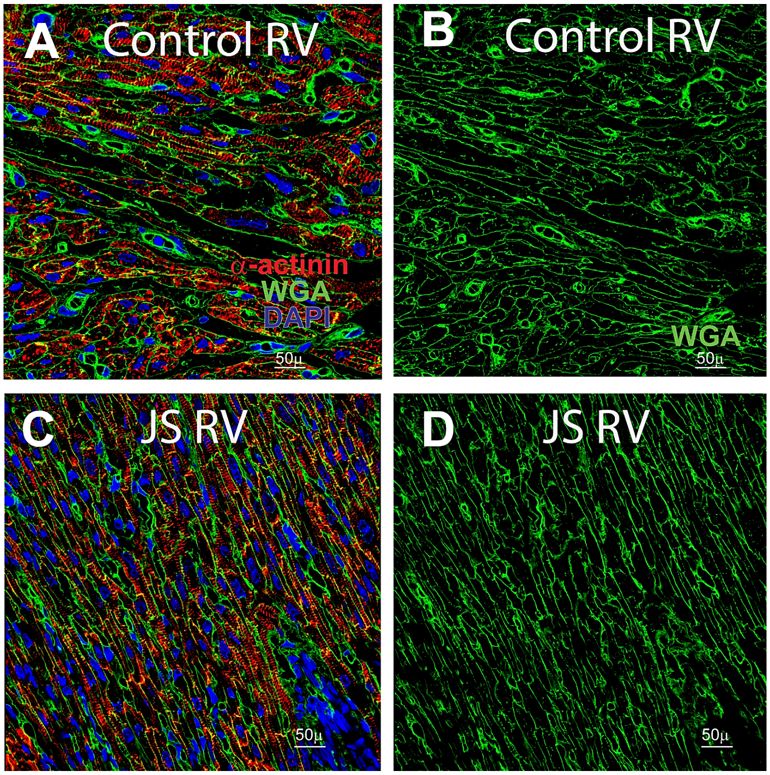


**Supplemental Figure 4: (Left)** Merged RV images from control and JS patients stained for α-actinin, WGA, and DAPI. **(Right)** WGA staining only, showing cell membranes.

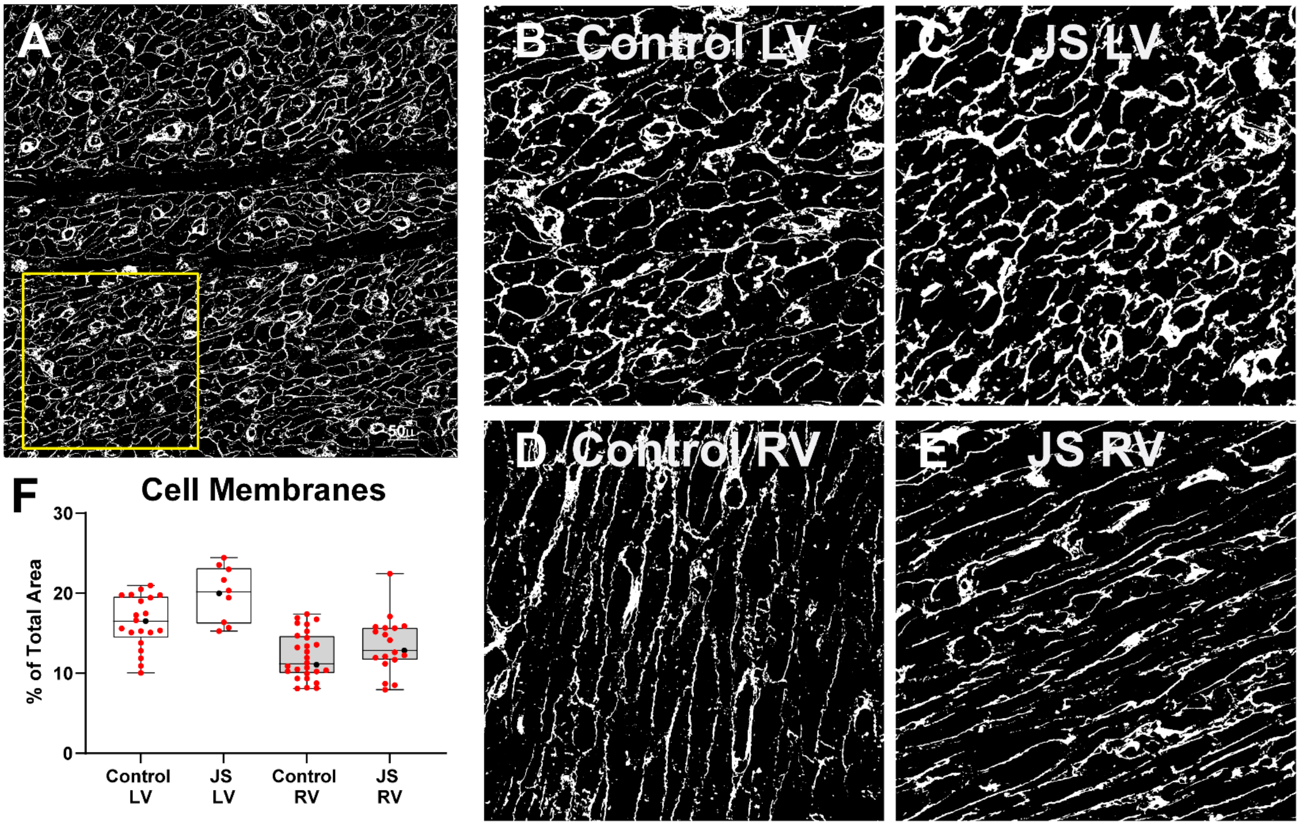


**Supplemental Figure 5: (A)** Binary images generated from a WGA-stained control LV. A 1500-pixel-square ROI (yellow box) was used to identify regions within stained sections containing myocardial cells. **(B-E)** Representative ROIs from control and JS ventricles. **(F)** Quantification of WGA-stained regions as a percentage of the total area. Black data points are values obtained for images in **(A-D)**. Plots show minimum, first quartile, median, third quartile, and maximum.

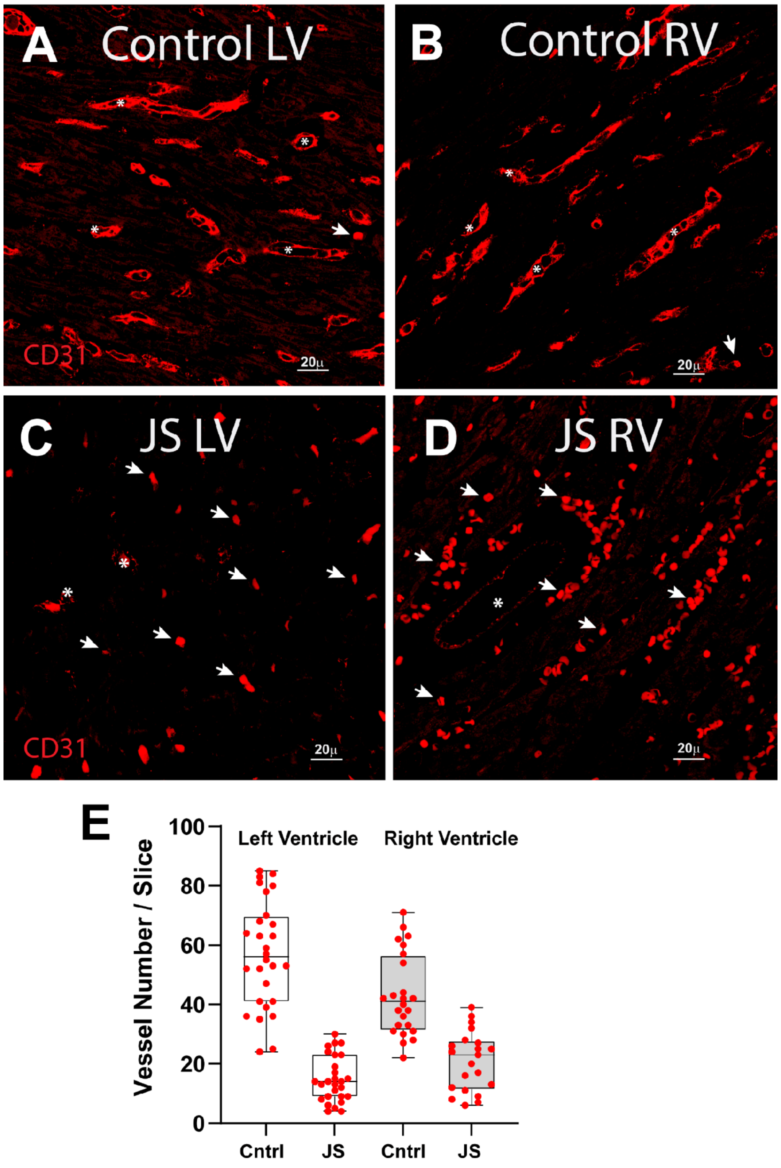


**Supplemental Figure 6:** CD31 immunoreactivity in **(A)** control left ventricle, **(B)** control right ventricle, **(C)** JS patient left ventricle, and **(D)** JS patient right ventricle. Acquisition settings were the same for all tissue samples. Arrows indicate examples of stained blood cells; * indicates examples of blood vessels. **(E)** Quantification of vessel number per tissue slice.

**Supplemental Movie Frog_1**. FLFM recording of wild type tadpole heart. Video was taken at 10ms intervals for ~7 seconds. XY, XZ, and YZ views were shown. Heart cells were labelled with nuclear GFP (H2b-GFP). Ventricle is to the center and outflow tract is to the bottom right.

**Supplemental Movie Frog_2**. FLFM recording of Ets1 KD tadpole heart. Video was taken at 10ms intervals for ~7 seconds. XY, XZ, and YZ views were shown. Heart cells were labelled

**Supplemental Movie Fish_3.** High speed optical recording of 72hpf zebrafish hearts from sham injected controls and ETS1, ETS2 and ETV2 triple KO fish. Anterior ventricle and outflow tract are to the left in each video.

**Supplemental Movie Fish_4.** High speed optical recording of 72hpf zebrafish hearts from sham injected controls and ETS1 KO fish. Anterior ventricle and outflow tract are to the left in each video.

#### Supplemental Table 1: Tadpole Heart Performance

| Performance Parameter | | Control (n=9) | Ets1-MO (n=7) | P-value |
|---|---|---|---|---|
| Heart Rate (per minute) | Min | 112 | 94 | 0.184698797 |
| | 1st Quartile | 121.2 | 117.8 | |
| | Median | 155 | 141 | |
| | 3rd Quartile | 159.5 | 141.8 | |
| | Max | 172 | 150 | |
| | Average | 144 | 129.285714 | |
| Systolic Ventricular Size ($mm^3$) | Min | 0.0059 | 0.0055 | 0.093789755 |
| | 1st Quartile | 0.008225 | 0.0079 | |
| | Median | 0.0144 | 0.0108 | |
| | 3rd Quartile | 0.02337 | 0.01115 | |
| | Max | 0.0251 | 0.0134 | |
| | Average | 0.0153165 | 0.00972791 | |
| Diastolic Ventricular Size ($mm^3$) | Min | 0.0125 | 0.0061 | 0.01400435 |
| | 1st Quartile | 0.01358 | 0.0098 | |
| | Median | 0.0244 | 0.0127 | |
| | 3rd Quartile | 0.03205 | 0.0156 | |
| | Max | 0.0356 | 0.0179 | |
| | Average | 0.02292091 | 0.01230797 | |
| Systolic Chamber Volume ($mm^3$) | Min | 0.0048 | 0.0036 | 0.100780704 |
| | 1st Quartile | 0.006125 | 0.006425 | |
| | Median | 0.0121 | 0.009 | |
| | 3rd Quartile | 0.02102 | 0.009525 | |
| | Max | 0.0238 | 0.0113 | |
| | Average | 0.01336204 | 0.00799324 | |
| Diastolic Chamber Volume ($mm^3$) | Min | 0.0103 | 0.0044 | 0.013364943 |
| | 1st Quartile | 0.01175 | 0.0082 | |
| | Median | 0.0209 | 0.011 | |
| | 3rd Quartile | 0.02832 | 0.01167 | |
| | Max | 0.034 | 0.0152 | |
| | Average | 0.0201836 | 0.01007899 | |
| Ejection Fraction | Min | 0.2123 | 0.128 | 0.002106312 |
| | 1st Quartile | 0.2747 | 0.1762 | |
| | Median | 0.4094 | 0.2036 | |
| | 3rd Quartile | 0.476 | 0.2378 | |
| | Max | 0.5299 | 0.257 | |
| | Average | 0.37793572 | 0.20267941 | |
| Cardiac Output ($mm^3$/minute) | Min | 0.7673 | 0.0772 | 5.02884E-07 |
| | 1st Quartile | 0.8488 | 0.1806 | |
| | Median | 0.9339 | 0.2352 | |
| | 3rd Quartile | 1.072 | 0.3792 | |
| | Max | 1.22 | 0.5503 | |
| | Average | 0.95768451 | 0.28091109 | |

**Supplemental Table 2: AVC and OFT development in zebrafish**

| | Identifiable AVC | Patent OFT |
|---|---|---|
| **Control** | 13/13 (100%) | 13/13 (100%) |
| **Ets1,2/Etv2 Crispant** | 9/17 (53%) | 2/17 (12%) |
| **Ets1 Crispant** | 18/20 (90%) | 18/20 (90%) |

**Supplemental Table 3: Qualitative assessment of α-actinin stained images for myofibrillar structure (blinded).**

| | Control LV | JS LV | Control RV | JS RV |
|---|---|---|---|---|
| **Organized** | 47% | 17% | 45% | 17% |
| **Disorganized** | 21% | 50% | 14% | 52% |
| **Both Types** | 32% | 33% | 41% | 31% |
| **N** | 17 | 9 | 22 | 21 |

### Supplemental Table 4: Fly Heart Performance

#### Adult Fly Heart Function: Pointed-RNAi

| Measure | Genotype | Mean | SD |
|---|---|---|---|
| End Diastolic Diameter (EDD) | Control (n=16) | 83.7 | 11.7 |
| | Pnt-RNAi (n=17) | 74.5 | 12.2 |
| *Unpaired t-test: t=2.214, df=31, p=0.0343 (*)* | | | |
| End Systolic Diameter (ESD) | Control | 51.7 | 9.4 |
| | Pnt-RNAi | 48.9 | 9.4 |
| *Unpaired t-test: t=0.8591, df=31, p=0.3969 (ns)* | | | |
| Fractional Shortening (FS) | Control | 0.38 | 0.04 |
| | Pnt-RNAi | 0.34 | 0.07 |
| *Unpaired t-test: t=2.188, df=31, p=0.0363 (*)* | | | |
| Cardiac Output (CO) | Control | 5315 | 1933 |
| | Pnt-RNAi | 3456 | 1703 |
| *Unpaired t-test: t=2.935, df=31, p=0.0062 (**)* | | | |
| Arrhythmicity Index (AI) | Control | 0.07 | 0.07 |
| | Pnt-RNAi | 0.10 | 0.07 |
| *Unpaired t-test: t=1.044, df=31, p=0.3045 (ns)* | | | |
| Heart Period (HP) | Control | 0.70 | 0.18 |
| | Pnt-RNAi | 0.81 | 0.22 |
| *Unpaired t-test: t=1.56, df=31, p=0.1288 (ns)* | | | |
| Diastolic Interval (DI) | Control | 0.46 | 0.18 |
| | Pnt-RNAi | 0.56 | 0.22 |
| *Unpaired t-test: t=1.421, df=31, p=0.1654 (ns)* | | | |
| Systolic Interval (SI) | Control | 0.23 | 0.02 |
| | Pnt-RNAi | 0.25 | 0.02 |
| *Unpaired t-test: t=1.778, df=31, p=0.0852 (ns)* | | | |

#### Adult Fly Heart Function: Pointed and Tinman Interaction

| Measure | Genotype | Mean | SD |
|---|---|---|---|
| End Diastolic Diameter (EDD) | w1118 (n=13) | 81.6 | 7.8 |
| | EC40/w1118 (n=17) | 79.9 | 8.6 |
| | pntD88/w1118 (n=14) | 85.2 | 8.9 |
| | pntD88/EC40 (n=14) | 50.8 | 15.6 |
| *Two-way ANOVA: EC40 p=<0.0001 (****) \| pntD88 p=<0.0001 (****) \| Interaction p=<0.0001 (****)* | | | |
| End Systolic Diameter (ESD) | w1118 | 51.1 | 5.3 |
| | EC40/w1118 | 49.1 | 6.7 |
| | pntD88/w1118 | 52.8 | 6.7 |
| | pntD88/EC40 | 36.8 | 11.5 |
| *Two-way ANOVA: EC40 p=<0.0001 (****) \| pntD88 p=0.014 (*) \| Interaction p=0.0014 (**)* | | | |
| Fractional Shortening (FS) | w1118 | 0.37 | 0.04 |
| | EC40/w1118 | 0.39 | 0.04 |
| | pntD88/w1118 | 0.38 | 0.06 |
| | pntD88/EC40 | 0.27 | 0.08 |
| *Two-way ANOVA: EC40 p=0.0028 (**) \| pntD88 p=0.0006 (***) \| Interaction p=0.0002 (***)* | | | |
| Cardiac Output (CO) | w1118 | 5179 | 1562 |
| | EC40/w1118 | 5106 | 1622 |
| | pntD88/w1118 | 3837 | 1093 |
| | pntD88/EC40 | 1239 | 760 |
| *Two-way ANOVA: EC40 p=0.0003 (***) \| pntD88 p=<0.0001 (****) \| Interaction p=0.0007 (***)* | | | |
| Arrhythmicity Index (AI) | w1118 | 0.50 | 1.09 |
| | EC40/w1118 | 0.24 | 0.21 |
| | pntD88/w1118 | 0.34 | 0.39 |
| | pntD88/EC40 | 0.30 | 0.33 |
| *Two-way ANOVA: EC40 p=0.994 (ns) \| pntD88 p=0.2509 (ns) \| Interaction p=0.659 (ns)* | | | |
| Heart Period (HP) | w1118 | 0.71 | 0.23 |
| | EC40/w1118 | 0.71 | 0.19 |
| | pntD88/w1118 | 1.05 | 0.29 |
| | pntD88/EC40 | 1.00 | 0.33 |
| *Two-way ANOVA: EC40 p=0.7415 (ns) \| pntD88 p=<0.0001 (****) \| Interaction p=0.7347 (ns)* | | | |
| Diastolic Interval (DI) | w1118 | 0.47 | 0.21 |
| | EC40/w1118 | 0.47 | 0.19 |
| | pntD88/w1118 | 0.81 | 0.28 |
| | pntD88/EC40 | 0.77 | 0.31 |
| *Two-way ANOVA: EC40 p=0.7694 (ns) \| pntD88 p=0.2826 (ns) \| Interaction p=0.7624 (ns)* | | | |
| Systolic Interval (SI) | w1118 | 0.24 | 0.03 |
| | EC40/w1118 | 0.24 | 0.02 |
| | pntD88/w1118 | 0.24 | 0.02 |
| | pntD88/EC40 | 0.24 | 0.03 |
| *Two-way ANOVA: EC40 p=0.5115 (ns) \| pntD88 p=0.6045 (ns) \| Interaction p=0.8342 (ns)* | | | |

### Supplemental Table 5: Fish Heart Performance

| | | | |
|---|---|---|---|
| Diastolic Surface Area | Control Atrium (n=14) | 5921.5 | 1131.2 |
| | ETS1 KO Atrium (n=19) | 6097.0 | 1256.7 |
| | Control Ventricle (n=14) | 5885.8 | 639.5 |
| | ETS1 KO Ventricle (n=18) | 6252.5 | 1347.7 |
| *One-way ANOVA: Atrium Control Vs. ETS1 KO p=0.772 (ns) \| Ventricle Control Vs. ETS1 KO p=0.9698 (ns)* | | | |
| Systolic Surface Area | Control Atrium (n=15) | 3531.7 | 698.5 |
| | ETS1 KO Atrium (n=20) | 4117.3 | 902.0 |
| | Control Ventricle (n=15) | 3698.4 | 466.7 |
| | ETS1 KO Ventricle (n=19) | 4496.6 | 821.8 |
| *One-way ANOVA: Atrium Control Vs. ETS1 KO p=0.0142 (*) \| Ventricle Control Vs. ETS1 KO p=0.0219 (*)* | | | |
| Fractional Area Change | Control Atrium (n=15) | 0.399 | 0.057 |
| | ETS1 KO Atrium (n=20) | 0.313 | 0.124 |
| | Control Ventricle (n=15) | 0.369 | 0.058 |
| | ETS1 KO Ventricle (n=19) | 0.272 | 0.098 |
| *One-way ANOVA: Atrium Control Vs. ETS1 KO p=0.0021 (**) \| Ventricle Control Vs. ETS1 KO p=0.0009 (***)* | | | |
| Heart Period | Control Atrium (n=13) | 0.455 | 0.017 |
| | ETS1/2/ETV2 KO Atrium (n=17) | 0.668 | 0.204 |
| | Control Ventricle (n=13) | 0.443 | 0.016 |
| | ETS1/2/ETV2 KO Ventricle (n=17) | 0.681 | 0.252 |
| *One-way ANOVA: Atrium Control Vs. ETS KO p=0.0031 (**) \| Ventricle Control Vs. ETS KO p=0.0009 (***)* | | | |
| End Diastolic | Control Atrium (n=13) | 11881.1 | 1800.6 |
| | ETS1/2/ETV2 KO Atrium (n=17) | 8252.7 | 2922.5 |
| | Control Ventricle (n=13) | 11079.8 | 1997.6 |
| | ETS1/2/ETV2 KO Ventricle (n=17) | 5627.5 | 1947.8 |
| *One-way ANOVA: Atrium Control Vs. ETS KO p<0.0001 (***) \| Ventricle Control Vs. ETS KO p<0.0001 (****)* | | | |
| End Systolic | Control Atrium (n=13) | 7346.9 | 1255.6 |
| | ETS1/2/ETV2 KO Atrium (n=17) | 6687.9 | 2423.3 |
| | Control Ventricle (n=13) | 6991.8 | 1241.3 |
| | ETS1/2/ETV2 KO Ventricle (n=17) | 4723.0 | 1579.8 |
| *One-way ANOVA: Atrium Control Vs. ETS KO p<0.5251 (ns) \| Ventricle Control Vs. ETS KO p<0.0017 (**)* | | | |
| FAC | Control Atrium (n=13) | 0.379 | 0.081 |
| | ETS1/2/ETV2 KO Atrium (n=17) | 0.183 | 0.077 |
| | Control Ventricle (n=13) | 0.366 | 0.063 |
| | ETS1/2/ETV2 KO Ventricle (n=17) | 0.142 | 0.089 |
| *One-way ANOVA: Atrium Control Vs. ETS KO p<0.0001 (****) \| Ventricle Control Vs. ETS KO p<0.0001 (****)* | | | |
| Cardiac Output (μ L/min) | Control (n=13) | 0.408 | 0.149 |
| | ETS1/2/ETV2 KO (n=17) | 0.068 | 0.068 |
| *Unpaired T-Test: Control Vs. ETS KO p<0.0001 (****)* | | | |

**Supplemental Table 6: Fly lines**

l[MI04868-GFSTF.1]

pnt[Δ88], pnt[MI03880]

svp[AE127-lacZ] all kept balanced over twi>>eGFP-labeled TM3 chromosome

*Twist*-GAL4

*Pnt*-RNAi- (VDRC KK105390; GD7171)

$Tin^{EC40}$

**Supplemental Table 7: Antibodies used**

anti-ß-galactosidase (mouse mab 40-1a from DSHB, rabbit polyclonal from Cappel)

anti-GFP (rabbit polyclonal from Rockland, mouse mab 3E6 from Life Technologies:A11120 )

anti-Doc3+2 (guinea pig polyclonal, Reim et al., 2003)

anti-Mef2 (rabbit polyclonal, from H.T. Nguyen)

*zfh-1* (gift of J. Skeath)

*odd-skipped* (Ward and Skeath, 2000)

*even-skipped* (Frasch et al., 1987)

*tinman* (gift of M. Frasch)

*seven-up* (DSHB clone 2D3)
